# HIV incidence among women engaging in sex work in sub-Saharan Africa: a systematic review and meta-analysis

**DOI:** 10.1101/2023.10.17.23297108

**Authors:** Harriet S Jones, Rebecca L Anderson, Henry Cust, R. Scott McClelland, Barbra A. Richardson, Harsha Thirumurthy, Kalonde Malama, Bernadette Hensen, Lucy Platt, Brian Rice, Frances M Cowan, Jeffrey W. Imai-Eaton, James R Hargreaves, Oliver Stevens

## Abstract

**Introduction:** HIV incidence among women in sub-Saharan Africa (SSA) has declined steadily, but it is unknown whether new infections among women who engage in sex work (WESW) have declined at a similar rate. We synthesised estimates of HIV incidence among WESW in SSA and compared these to the wider female population to understand levels and trends in incidence over time.

**Methods:** We searched Medline, Embase, Global Health, Popline, Web of Science, and Google Scholar from January 1990 to October 2022, and grey literature for estimates of HIV incidence among WESW in SSA. We included studies reporting empirical estimates in any SSA country. We calculated incidence rate ratios (IRR) compared to age-district-year matched total female population incidence estimates. We conducted a meta-analysis of IRRs and used a continuous mixed-effects model to estimate changes in IRR over time.

**Results:** From 32 studies between 1985 and 2020, 2,194 new HIV infections were observed in WESW over 51,000 person-years (py). Median HIV incidence was 4.3/100py (IQR 2.8-7.0/100py), declining from a median of 5.96/100py between 1985 and 1995 to a median of 3.2/100py between 2010 and 2020. Incidence among WESW was nine times higher than in matched total population women (RR 8.6, 95%CI: 5.7-12.9), and greater in Western and Central Africa (RR 22.4, 95%CI: 11.3-44.3) than in Eastern and Southern Africa (RR 5.3, 95%CI: 3.7–7.6). Annual changes in log IRRs were minimal (–0.1% 95%CI: −6.9 to +6.8%).

**Conclusions:** Across SSA, HIV incidence among WESW remains disproportionately high compared to the total female population but showed similar rates of decline between 1990 and 2020. Improved surveillance and standardisation of approaches to obtain empirical estimates of sex worker incidence would enable a clearer understanding of whether we are on track to meet global targets for this population and better support data-driven HIV prevention programming.

## Introduction

New HIV infections have steadily declined among women in sub-Saharan Africa (SSA) since 1994, including by 65% between 2010–2022 in Eastern and Southern Africa (ESA).^1^ However, among women across SSA, those who engage in sex work bear a disproportionate burden of HIV.^1, 2^ Women who engage in sex work (WESW) comprise an estimated 1.2% of woman aged 15-49 in SSA, but an estimated 3.5% of women living with HIV in the region.^2^ HIV prevention programmes for WESW were a core component of the early HIV response in SSA, however reduced funding and a shift to general population programming meant a decline in programmes focused on WESW from the early 2000s.^3^ More recently, renewed energy in targeted approaches for key populations has resulted in the re-expansion of programming for WESW.^4–, 6^ Assessing the success of these efforts on preventing new HIV infections is challenging, and it is unknown whether HIV incidence among WESW has declined in line with women in the wider population.

Despite increasing surveillance and programmatic support for WESW, studies of HIV incidence in SSA remain infrequent and challenging. Identifying and following-up WESW is often difficult due to the heterogeneous, informal, and hidden nature of sex work, commonly driven by stigma and criminalisation.^7^ For these reasons WESW are largely unidentified in population-based surveys and constructing a national sampling frame usually impractical. Sex work encompasses a broad spectrum of sexual transactions occurring in a range of settings, from on the streets, to in homes, brothels, or hotels.^8^ Women who engage in transactional sex may not self-identify as sex workers. For surveillance, this presents challenges, as those not identifying as sex workers are unlikely to present at WESW-dedicated programmes. Additionally, mobility among WESW is high,^9^ and repeated initiation and cessation of sex work common, thus programmes and cohort studies encounter challenges with loss to follow-up.^10, 11^ To mitigate these challenges, there is growing focus on techniques leveraging cross-sectional data to estimate incidence,^12, 13^ whilst recruitment approaches such as respondent-driven sampling (RDS) and time-location sampling (TLS) are increasingly used to capture more representative samples of WESW.

As countries seek to reach the global goal of ending AIDS as a public health threat by 2030 through ending inequalities, quantifying HIV incidence trends in WESW is required to guide national HIV programme planning and delivery. In this study we aimed to synthesise and appraise empirical estimates of HIV incidence in WESW in SSA, estimate relative HIV incidence between WESW and the total female population for WCA and ECA, and estimate the change in relative HIV incidence over time.

## Methods

We searched published and grey literature to identify empirical estimates of HIV incidence among WESW in SSA. We conducted two searches of peer-reviewed literature available in English and published between January 1990 and December 2022. Medline, EMBASE, Popline, Web of Science, and Global Health were searched on 19^th^ June 2019 using medical subject headings (MeSH) and text words adapted for each database covering three domains: ‘Female sex workers’, ‘HIV’, and ‘sub-Saharan Africa’. Searches were updated in January 2023 in Medline, EMBASE, Global Health, and Google Scholar using text words addressing four domains: ‘sex workers’, ‘sub-Saharan Africa’, ‘HIV’ and ‘incidence’ (Supplementary Text S1). The initial title, abstract, and full text screening was conducted by HSJ and HC in 2019 for an earlier review of HIV testing among WESW^14^ and full texts subsequently searched for those reporting HIV incidence. A second search was conducted by RLA in January 2023, with all papers selected for final inclusion based on consensus between HSJ, RLA, and OS. RLA additionally searched key population biobehavioural surveillance reports, collated during an earlier key population data collation exercise.^2^

### Study inclusion

For inclusion, studies needed to report an empirically measured estimate of HIV-test confirmed HIV incidence in WESW in any SSA country or report the number of HIV events and total person-years (py) at-risk to manually calculate incidence. WESW could be women either self-reporting being a sex worker, engaged in a sex worker programme, or reporting selling or exchanging sex for money or goods. For multi-country studies, nationally disaggregated data were required. For closed cohort studies reporting annual estimates, only the overall estimate of incidence from the study was extracted to mitigate against artificial declines in incidence observed with follow-up of the same individuals. Studies were excluded if the study cohort was exposed to an intervention specifically thought to have impacted HIV incidence, i.e. HIV pre-exposure prophylaxis (PrEP). In randomised control trials (RCT), data from the study control arm were included and, in interventional cohort studies, data from early in the intervention or pre-intervention period included. Conference abstracts or published short communications were excluded. Where multiple papers reported incidence estimates for the same study population, the paper reporting the greatest number of person-years of follow-up was selected as the primary study for data extraction (Figure 1).

**Figure 1:**
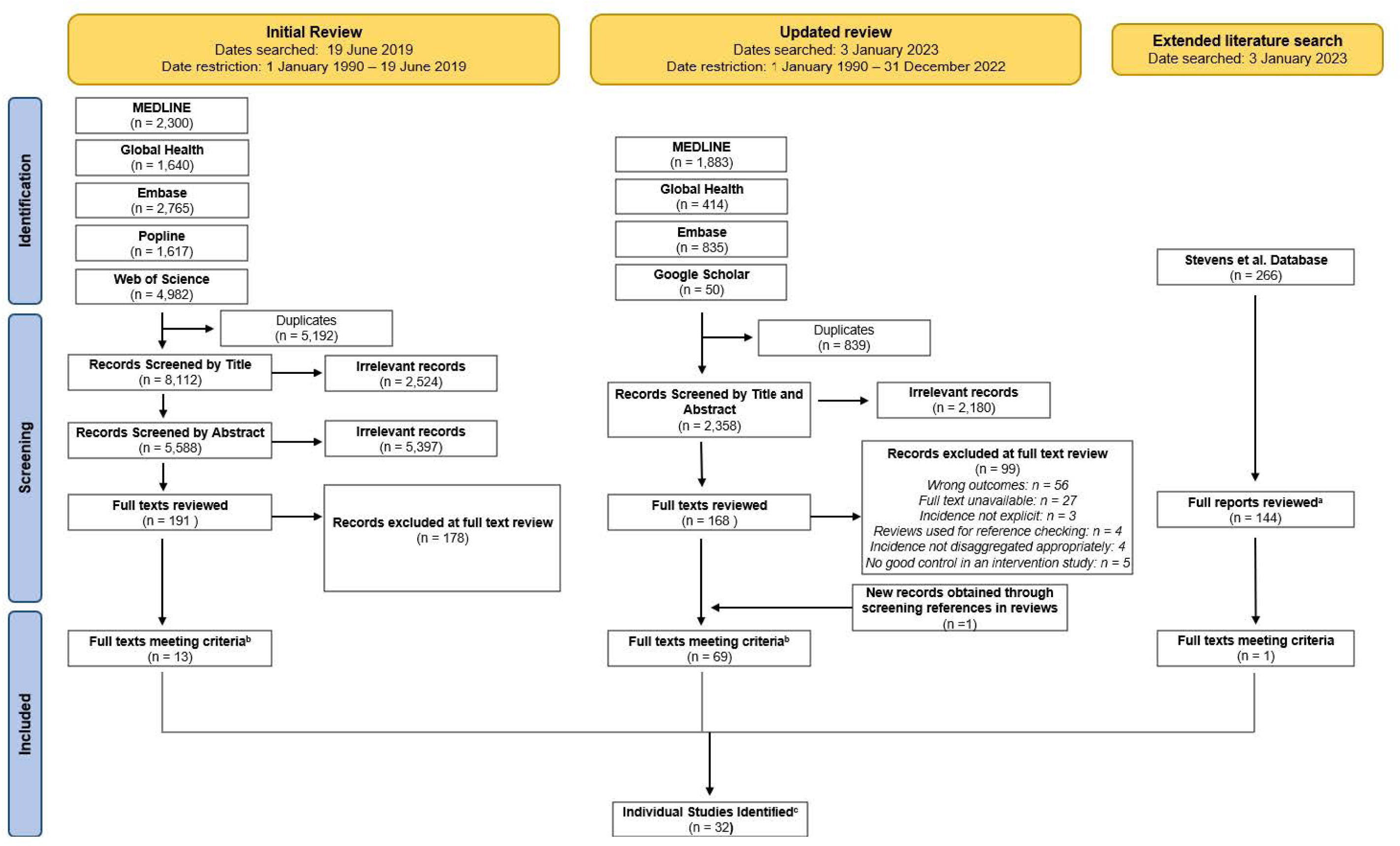
Flow diagram summarising identification, screening, and inclusion of studies. a: Reports retrieved through key term search for the word “incidence” in File Explorer. b: **Full texts meeting criteria** include multiple papers reporting on the same study. c. **Individual Studies Identified** are the primary texts which data were extracted from.

### Data extraction

From each study, two authors extracted the study location (country, subnational area), time period, study population definition, recruitment strategy, mean/median age and recruitment range, incidence measurement method (e.g. recency testing, repeat HIV tests), study type (e.g. cohort, RCT), sample size, person-years of follow-up, number of seroconversions, incidence per 100py, and confidence intervals, disaggregated by age-group where available. We contacted study authors where estimates were not clearly available in existing publications (n=1),^15^ not disaggregated to WESW (n=1),^16^ or from studies reporting data for extended time periods which could provide annual estimates to assess temporal trends (n=2).^17, 18^

### Quality assessment

We appraised studies using the Global HIV Quality Assessment Tool for Data Generated through Non-Probability Sampling (GHQAT).^19^ The tool comprises three domains: study design, study implementation and a measurement specific domain for HIV incidence. Domain specific assessment criteria are in the tool and supplementary material (Supplementary Table 2). We defined scoring requirements for subjective assessment criteria, stipulating reporting requirements needed to score positively. Measurement quality scored positively if studies stated their approach to seroconversion date measurement. Sample recruitment scored positively if studies used a network sampling approach (e.g. RDS^20^), TLS, or clinic sampling with peer recruitment. Where physical locations were used for recruitment, location mapping needed to be reported. Studies were also scored positively if study length was at minimum 12 months, retention >70%, tracing was conducted for participants lost-to-follow-up or statistical adjustments made to account for loss-to-follow-up. We chose not to score studies on overall reporting quality. Based on a score of 1 if studies met the assessment criteria and 0 if they did not, each was classified as good, fair, or poor for each domain, and then overall. Studies scoring ≥ 70% were classified as good, between 30% and 70% fair and <30% poor. We adapted scoring to account for exclusion of reporting criteria and for studies calculating incidence from cross-sectional recent infection testing or serial HIV prevalence measures.^12, 13^

### Data synthesis and meta-analysis

HIV incidence observations among WESW were matched to HIV incidence in the total female population by district (subnational location where each study was conducted), age, and year. Estimates missing age information were matched to incidence in total population women aged 15-39, reflecting the age distribution observed in sex workers in studies reporting age data. District-level estimates in 2022 for the total female population were extracted from UNAIDS subnational HIV estimates created using the Naomi model.^21^ Naomi is a district-level small-area estimation model that calibrates to nationally representative household survey and routine ART and antenatal health system data. District-level incidence estimates for 1985-2021 were created by extrapolating 2022 estimates backwards in time, parallel to UNAIDS national-level female incidence trajectories, assuming the proportional change in incidence at district-level mirrored that at national-level.^22, 23^ For studies where annual incidence estimates were unavailable, the midpoint study year was used for matching. When a subnational location was not specified in the text (n=3), national total population incidence was used as the comparator. One study presented incidence for a control group of women not engaged in sex work,^24^ which was used as the total population comparator instead of matching to UNAIDS’ district estimate.

We assessed correlation between WESW and matched total population female incidence and calculated incidence rate ratios (IRRs). We used a meta-analysis of IRRs, with study-country-district nested random effects to account for variation in study type, and regional fixed effects to stratify our pooled IRRs by region (ESA and Western and Central Africa (WCA)). We examined trends in IRRs over time using a Bayesian mixed-effects log-linear model, predicting log-IRRs using calendar year fixed effects, study-level random effects, and total female population incidence and person-years of follow-up as offsets (Supplementary Text S2). Lastly, we conducted case studies for the two countries with the most available data. We used data from an open cohort in Mombasa, Kenya,^18^ and from a national key populations programme in Zimbabwe run by the Centre for Sexual Health and HIV/AIDS Research Zimbabwe (CeSHHAR),^17^ to descriptively assess temporal trends between WESW and total female population incidence.

We conducted several sensitivity analyses. Firstly, to address uncertainty surrounding the district-level total population incidence estimates, we repeated the meta-analysis using national age-sex matched population incidence as the denominator for IRRs. To assess the impact of study quality, we repeated the meta-analysis filtered to only higher quality studies (those scoring 60% or above on the GHQAT). Finally, as the majority of empirical incidence estimates for WESW were from populations in Kenya and Zimbabwe, we refit the mixed-effects model to data from both countries. Analyses were implemented in R v4.2.1^25^ using the *metafor* v3.8.1^26^ and *R-INLA* v22.5.7^27^ packages.

### Ethical approval

This study received ethical approval from the Imperial College Research and Ethics Committee (#6412027). For use of CeSHHAR’s Key Populations programme data, ethical approval was obtained from the London School of Hygiene and Tropical Medicine (16543) and the Medical Research Council of Zimbabwe (MRCZ/A/2624).

## Results

We extracted 83 estimates of HIV incidence among WESW in sub-Saharan Africa from 32 studies reported in 69 peer-reviewed papers and one surveillance report (Tables 1 and 2). The majority of estimates were from ESA (78%; 65/83), predominantly from Kenya and Zimbabwe (59%, 49/83). In WCA, 18 incidence estimates were reported from eight countries. Median study year was 2008 (IQR: 2000-2015). Between 1985 and 2020, 2,194 new HIV infections were observed from 51,000py with median HIV incidence of 4.3/100py (interquartile range [IQR] 2.8-7.0/100py).

**Table 1:** Annual incidence estimates for women who engage in sex work (WESW) and total population women aged 15-39.

| Study<sup>a</sup> | Country | Study Period | Area<sup>b</sup> | Mid-point Year | Age Group<sup>c</sup> | WESW New HIV Infections | Person-years (py) of follow-up | WESW Incidence/100py | Total female population incidence/100/py | IRR<sup>d</sup> |
| --- | --- | --- | --- | --- | --- | --- | --- | --- | --- | --- |
| Ali (2020)<sup>e</sup> | Zimbabwe | 2011-2017 | Zimbabwe | 2014 | 15-24 | 7 | 105 | 6.3 | 1.12 | 5.64 |
|  |  |  |  |  | 25-39 | 6 | 175 | 3.3 | 0.92 | 3.58 |
| Botswana MoH (2013)<sup>f</sup> | Botswana | 2012 | Gaborone, Francistown, Kasane | 2012 | 15-39 | 46 | 450 | 12.5 (7.3, 17.1) | 1.65 | 7.57 |
| *Project Ubuzima* Braunstein (2011) | Rwanda | 2006-2008 | Kigali | 2007 | 15-39 | 3 | 91 | 3.3 (0.0, 7.0) | 0.47 | 6.96 |
| Chabata (2021) | Zimbabwe | 2017-2019 | Chinhoyi | 2018 | 15-24 | 16 | 226 | 7.1 (4.3, 11.5) | 0.51 | 13.83 |
|  |  |  | Karoi |  |  | 11 | 193 | 5.7 (3.2, 10.3) | 0.48 | 11.96 |
|  |  |  | Kwekwe |  |  | 12 | 278 | 4.3 (2.5, 7.6) | 0.71 | 6.04 |
|  |  |  | Zvishavane |  |  | 9 | 210 | 4.3 (2.2, 8.2) | 0.68 | 6.28 |
| Chersich (2014) | Kenya | 2006-2007 | Mombasa (Kisauni and Chaani) | 2007 | 15-39 | 10 | 381 | 2.6 | 0.58 | 4.54 |
| Diabate (2018) | Benin | 2008-2012 | Cotonou | 2010 | 15-39 | 6 | 425 | 1.4 (0.3, 2.5) | 0.28 | 5 |
| Faini (2022) | Tanzania | 2018-2019 | Dar es Salaam | 2019 | 15-24 | 12 | 278 | 4.3 | 0.05 | 89.86 |
|  |  |  |  |  | 25-34 | 5 | 234 | 2.1 | 0.06 | 36.35 |
|  |  |  |  |  | 35-44 | 4 | 97 | 4.1 | 0.05 | 88.44 |
| Forbi (2011) | Nigeria | 2006 | Nasarawa State | 2006 | 15-34 | 71 | 590 | 11.97 (8.5, 15.4) | 0.3 | 39.56 |
| Fowke (1996) | Kenya | 1985-1994 | Nairobi | 1990 | 15-39 | 239 | 569 | 42.0 | 3.01 | 13.96 |
| Ghys (2001) | Cote d'Ivoire | 1994-1998 | Abidjan | 1996 | 15-39 | 11 | 68 | 16.3 | 1.79 | 9.11 |
| Gilbert (2003) | Senegal | 1985-1999 | Dakar | 1987 | 15-24 | 34 | 1628 | 2.1 | 0.01 | 219.9 |
|  |  |  | Dakar |  | 25-34 | 91 | 5474 | 1.7 | 0.02 | 77.08 |
|  |  |  | Dakar |  | 35-44 | 58 | 1939 | 3.0 | 0.01 | 201.7 |
|  |  |  | Dakar |  | 45-49 | 12 | 260 | 4.6 | 0.01 | 597 |
| Jones (2023)<sup>g</sup> | Zimbabwe | 2010-2019 | Zimbabwe | 2010 | 15-39 | 3 | 56 | 5.4 (3.2, 10.9) | 1.33 | 4.04 |
|  |  |  |  | 2011 |  | 6 | 139 | 4.3 (2.2, 14.6) | 1.25 | 3.45 |
|  |  |  |  | 2012 |  | 15 | 264 | 5.7 (1.9, 22.8) | 1.16 | 4.88 |
|  |  |  |  | 2013 |  | 12 | 410 | 2.9 (1.5, 8.1) | 1.09 | 2.69 |
|  |  |  |  | 2014 |  | 33 | 731 | 4.5 (2.5, 8.2) | 1.01 | 4.46 |
|  |  |  |  | 2015 |  | 62 | 1402 | 4.4 (3.6, 5.4) | 0.93 | 4.78 |
|  |  |  |  | 2016 |  | 58 | 1857 | 3.1 (2.5, 4.1) | 0.82 | 3.81 |
|  |  |  |  | 2017 |  | 94 | 2422 | 3.9 (3.3, 4.65) | 0.71 | 5.46 |
|  |  |  |  | 2018 |  | 114 | 2946 | 3.9 (3.3, 4.7) | 0.57 | 6.75 |
|  |  |  |  | 2019 |  | 44 | 1432 | 3.1 (2.1, 5.0) | 0.49 | 6.32 |
| Kasamba (2019) | Uganda | Cohort 1: 2008-2017 | Kampala | 2010 | 15-39 | 59 | 2007 | 2.9 | 1.06 | 2.74 |
|  |  | Cohort 2: 2013-2017 |  | 2014 |  | 46 | 1394 | 3.3 | 0.86 | 3.82 |
|  |  |  |  | 2016 |  | 65 | 2138 | 3.0 | 0.77 | 3.89 |
| Kassanje (2022)<sup>f</sup> | South Africa | 2019 | South Africa | 2019 | 15-49 | 190 | 4130 | 4.6 (1.5, 8.5) | 1.1 | 4.18 |
| Kaul (2004) | Kenya | 1998-2002 | Nairobi | 2000 | 15-39 | 16 | 495 | 3.2 | 1.2 | 2.67 |
| Kerrigan (2017) | Tanzania | 2015-2017 | Mafinga | 2002 | 15-39 | 10 | 144 | 6.9 | 2.28 | 3.04 |
| Kilburn (2018) | South Africa | 2012-2015 | Mpumalanga | 2014 | 15-19 | 41 | 1273 | 3.2 | 1.85 | 1.74 |
| Laga (1994) | DRC | 1988-1991 | Kinshasa | 1988 | 15-39 | 70 | 880 | 11.7 | 0.32 | 36.55 |
| Lyons (2020) | Senegal | 2015-2017 | Dakar, Mbour, Theis | 2016 | 15-39 | 4 | 303 | 1.3 (0.5, 3.5) | 0.01 | 151.2 |
| Malama (2022) | Zambia | 2012-2017 | Ndola, Lusaka | 2015 | 15-44 | 24 | 884 | 2.7 | 1.59 | 1.73 |
| McClelland (2015)<sup>g</sup> | Kenya | 1993-2017 | Mombasa | 1993 | 15-39 | 20 | 335 | 6.0 | 3.02 | 1.97 |
|  |  |  |  | 1994 |  | 36 | 228 | 15.8 | 2.72 | 5.83 |
|  |  |  |  | 1995 |  | 24 | 213 | 11.3 | 2.29 | 4.92 |
|  |  |  |  | 1996 |  | 29 | 325 | 8.9 | 1.86 | 4.8 |
|  |  |  |  | 1997 |  | 41 | 364 | 11.3 | 1.56 | 7.24 |
|  |  |  |  | 1998 |  | 31 | 271 | 11.4 | 1.3 | 8.78 |
|  |  |  |  | 1999 |  | 24 | 207 | 11.6 | 1.13 | 10.28 |
|  |  |  |  | 2000 |  | 16 | 188 | 8.5 | 0.99 | 8.65 |
|  |  |  |  | 2001 |  | 8 | 216 | 3.7 | 0.88 | 4.22 |
|  |  |  |  | 2002 |  | 19 | 251 | 7.6 | 0.81 | 9.34 |
|  |  |  |  | 2003 |  | 14 | 245 | 5.7 | 0.73 | 7.8 |
|  |  |  |  | 2004 |  | 13 | 263 | 5.0 | 0.69 | 7.22 |
|  |  |  |  | 2005 |  | 14 | 271 | 5.2 | 0.65 | 7.98 |
|  |  |  |  | 2006 |  | 8 | 228 | 3.5 | 0.6 | 5.8 |
|  |  |  |  | 2007 |  | 7 | 244 | 2.9 | 0.58 | 4.98 |
|  |  |  |  | 2008 |  | 4 | 211 | 1.9 | 0.54 | 3.51 |
|  |  |  |  | 2009 |  | 9 | 163 | 5.5 | 0.52 | 10.73 |
|  |  |  |  | 2010 |  | 3 | 187 | 1.6 | 0.48 | 3.34 |
|  |  |  |  | 2011 |  | 3 | 283 | 1.1 | 0.45 | 2.37 |
|  |  |  |  | 2012 |  | 2 | 334 | 0.6 | 0.41 | 1.47 |
|  |  |  |  | 2013 |  | 4 | 280 | 1.4 | 0.37 | 3.84 |
|  |  |  |  | 2014 |  | 5 | 225 | 2.2 | 0.34 | 6.57 |
|  |  |  |  | 2015 |  | 5 | 222 | 2.3 | 0.31 | 7.25 |
|  |  |  |  | 2016 |  | 4 | 216 | 1.9 | 0.27 | 6.78 |
|  |  |  |  | 2017 |  | 3 | 237 | 1.3 | 0.24 | 5.27 |
| McKinnon (2015) | Kenya | 2008-2011 | Nairobi | 2010 | 15-39 | 34 | 1514 | 2.2 (1.6, 3.1) | 0.58 | 3.78 |
| Nagot (2005) | Burkina Faso | 1998-2002 | Bobo-Dioulasso | 2000 | 15-39 | 19 | 594 | 3.2 (1.9, 4.9) | 0.18 | 17.72 |
| Naiker (2015) | South Africa | 2004-2006 | Durban | 2005 | 25-49 | 10 | 248 | 4.0 | 1.82 | 2.22 |
|  |  |  |  |  | 15-24 | 8 | 59 | 13.5 | 2.53 | 5.32 |
| Nouaman (2022) | Cote d'Ivoire | 2016-2017 | San Pedro | 2017 | 15-39 | 4 | 188 | 3.3 | 0.08 | 43.75 |
|  |  |  | Abidjan |  |  | 3 | 293 | 1.6 | 0.14 | 11.08 |
| Price (2012) | Kenya | 2008 | Kilifi | 2007 | 15-39 | 9 | 339 | 2.7 (1.4, 5.1) | 0.36 | 7.51 |
|  |  |  | Nairobi |  |  | 2 | 527 | 0.4 (0.1, 1.5) | 0.7 | 0.57 |
| Priddy (2011) | Kenya | 2008 | Nairobi (Mukuru District) | 2008 | 15-39 | 5 | 89 | 5.6 (1.6, 12.0) | 0.66 | 8.54 |
| Riedner (2006) | Tanzania | 2000-2003 | Mbeya Region | 2002 | 15-39 | 19 | 99 | 19.2 | 1.32 | 14.56 |
| Roddy (1998) | Cameroon | 1994-1996 | Yaounde, Douala | 1995 | 15-39 | 46 | 698 | 6.6 | 0.7 | 9.4 |
| Thirumurthy<br>(2021)<sup>g</sup> | Kenya | 2017-2020 | Siaya County | 2018 | 15-39 | 0 | 939 | 0.0 | 0.41 | 0 |
| Van Damme<br>(2002) | South<br>Africa | 1996-2000 | KwaZulu-Natal | 1998 | 15-39 | 30 | 182 | 16.5 | 2.96 | 5.57 |
|  | Benin |  | Cotonou |  |  | 10 | 121 | 8.3 | 0.77 | 10.79 |
|  | Cote<br>d'Ivoire |  | Abidjan |  |  | 5 | 67 | 7.4 | 1.4 | 5.29 |
| Van der Loeff<br>(2001) | Guinea-Bissau | 1989-1998 | Guinea-Bissau | 1994 | 15-39 | 3 | 126 | 2.4 | 0.41 | 5.78 |
|  |  |  |  |  | 40-49 | 7 | 160 | 4.4 | 0.22 | 19.89 |
|  |  |  |  |  | 50-59 | 5 | 67 | 7.5 | 0.12 | 60.77 |

**Table 2:**
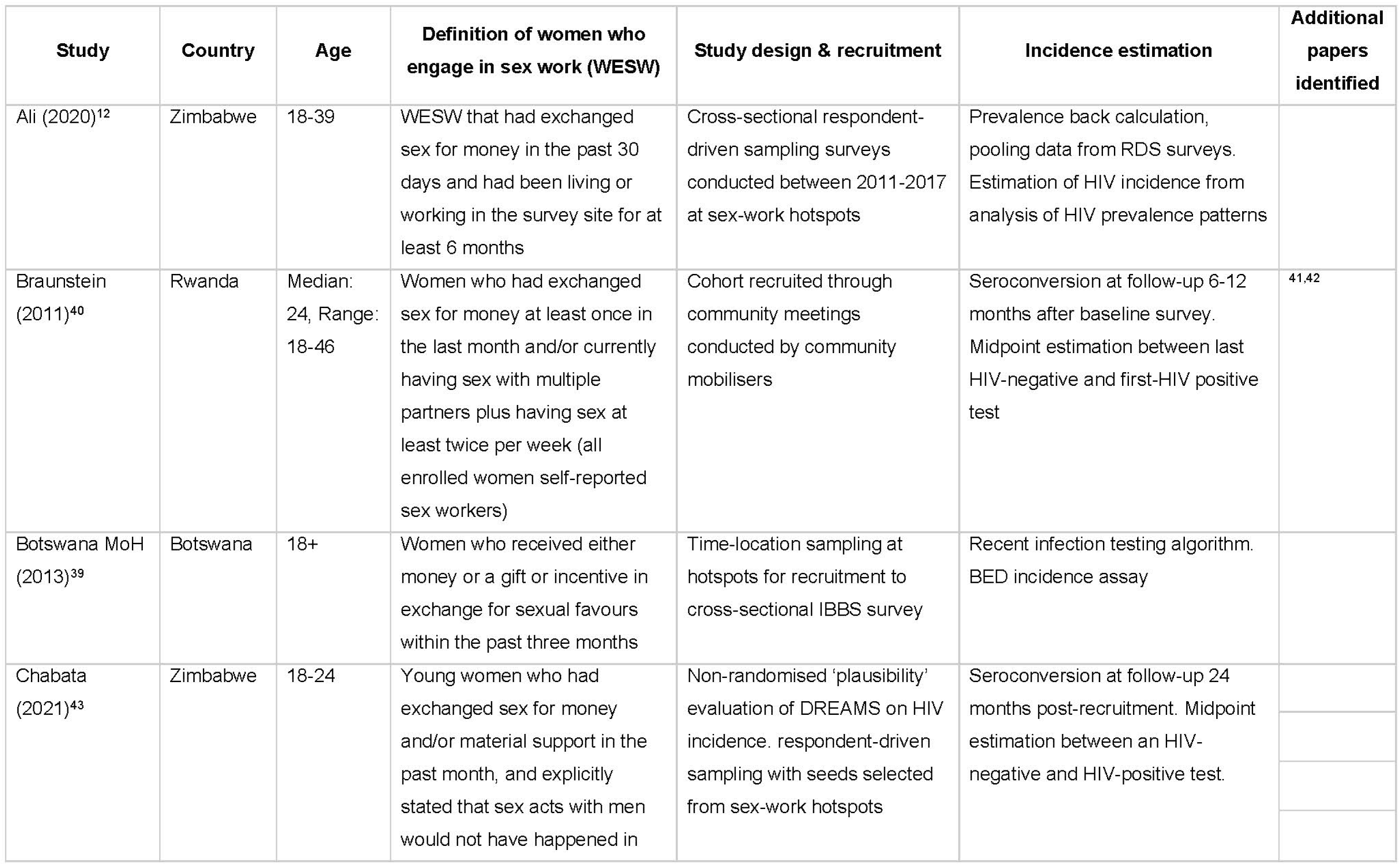
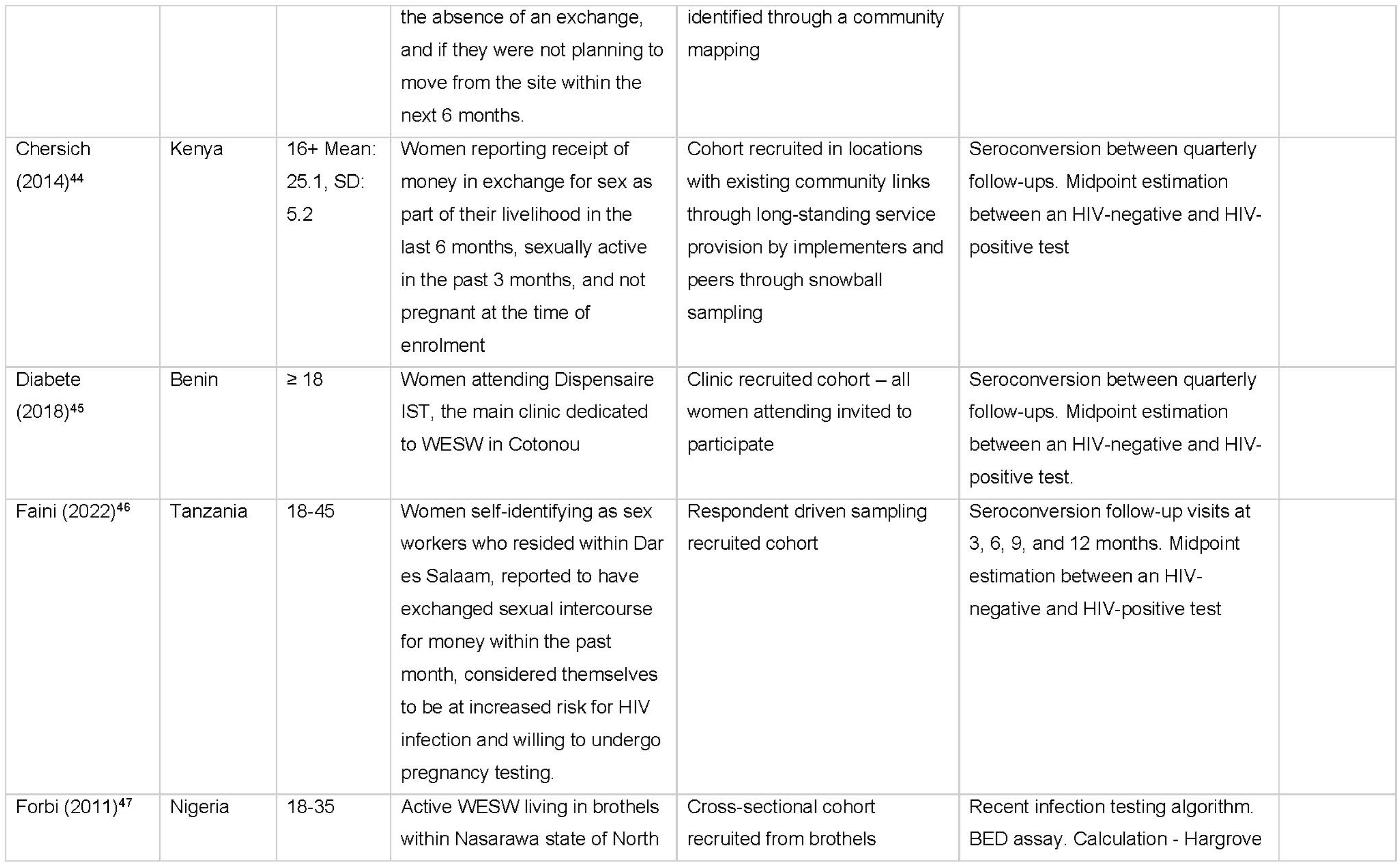
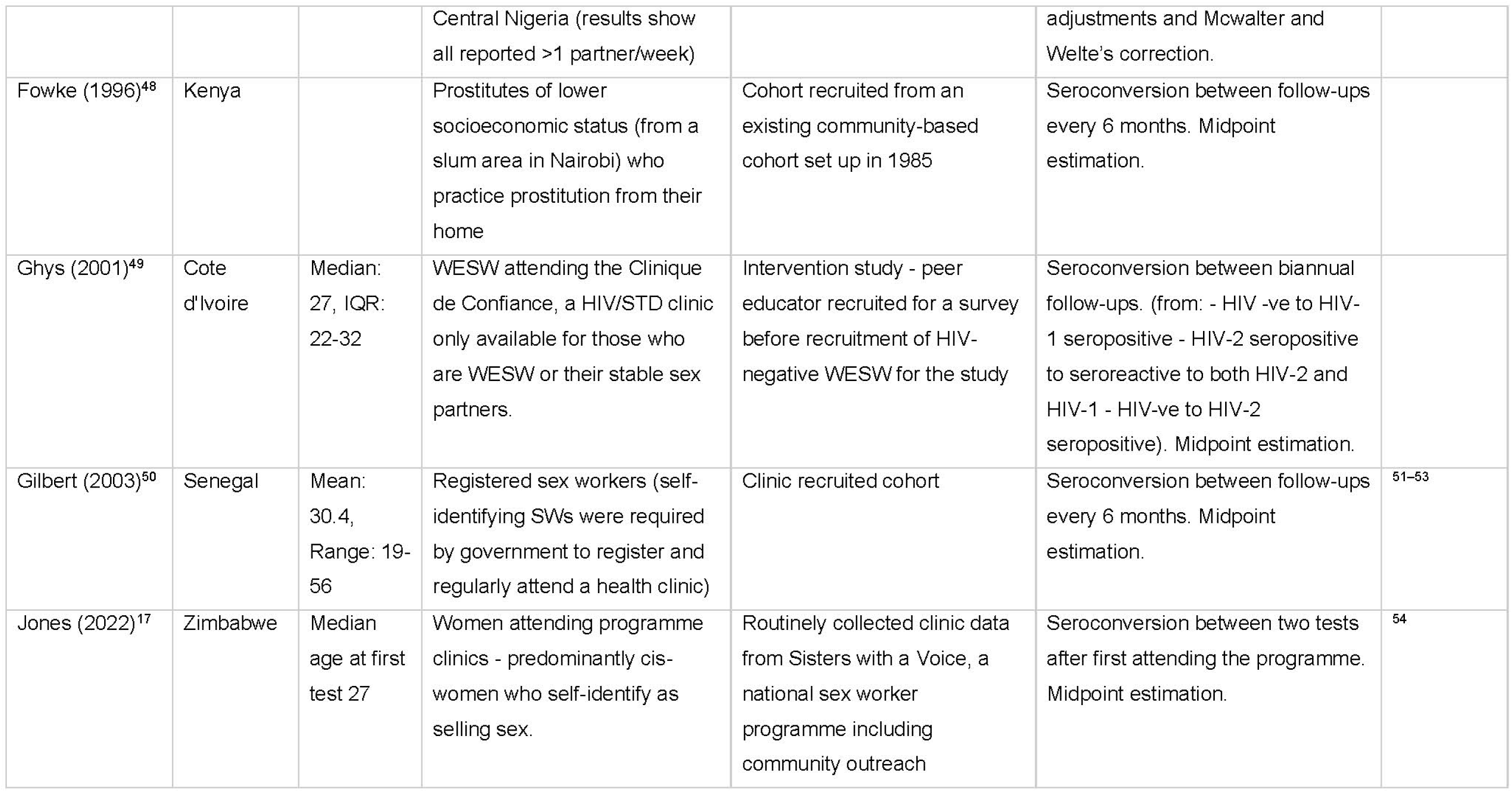
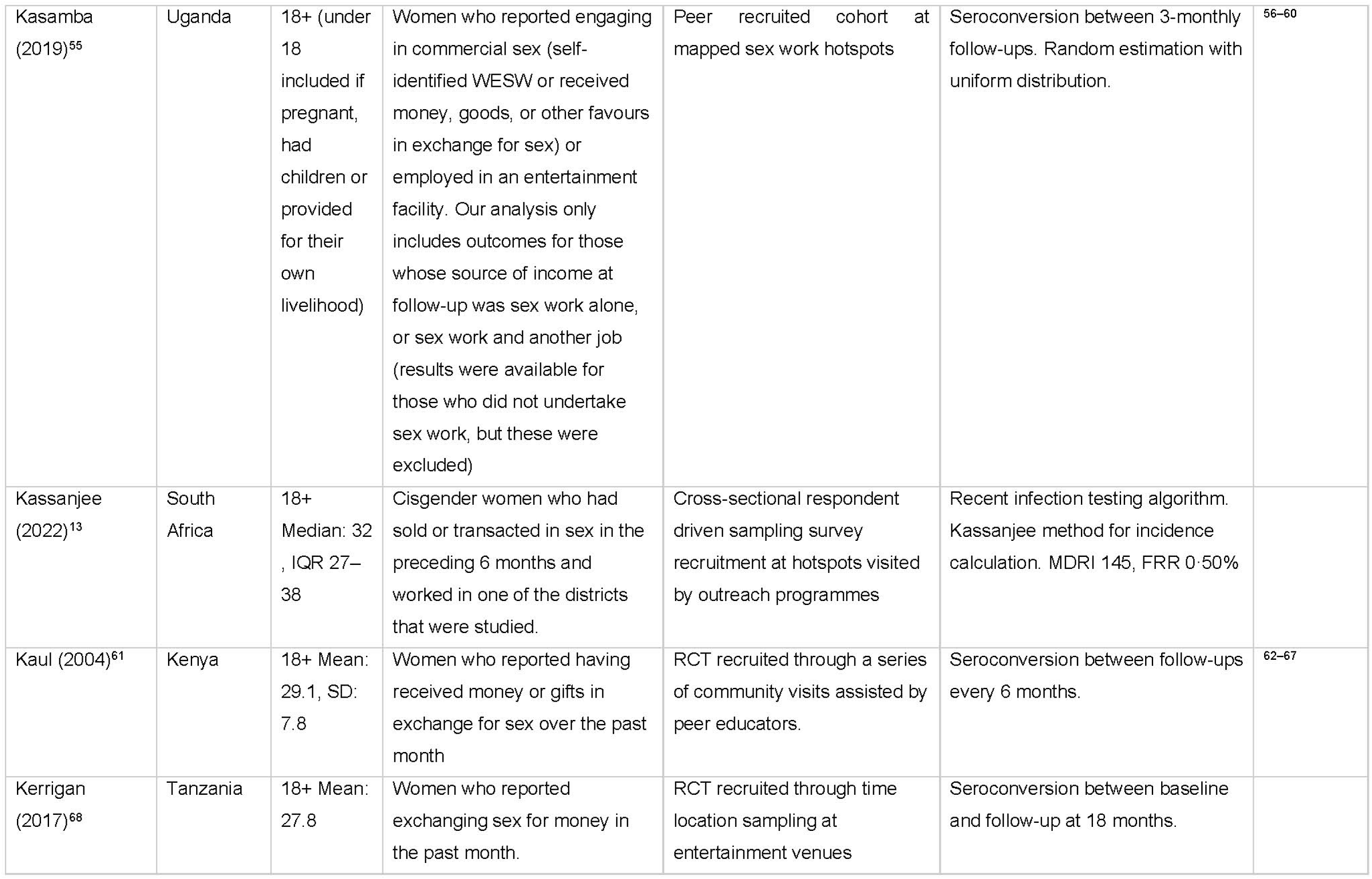
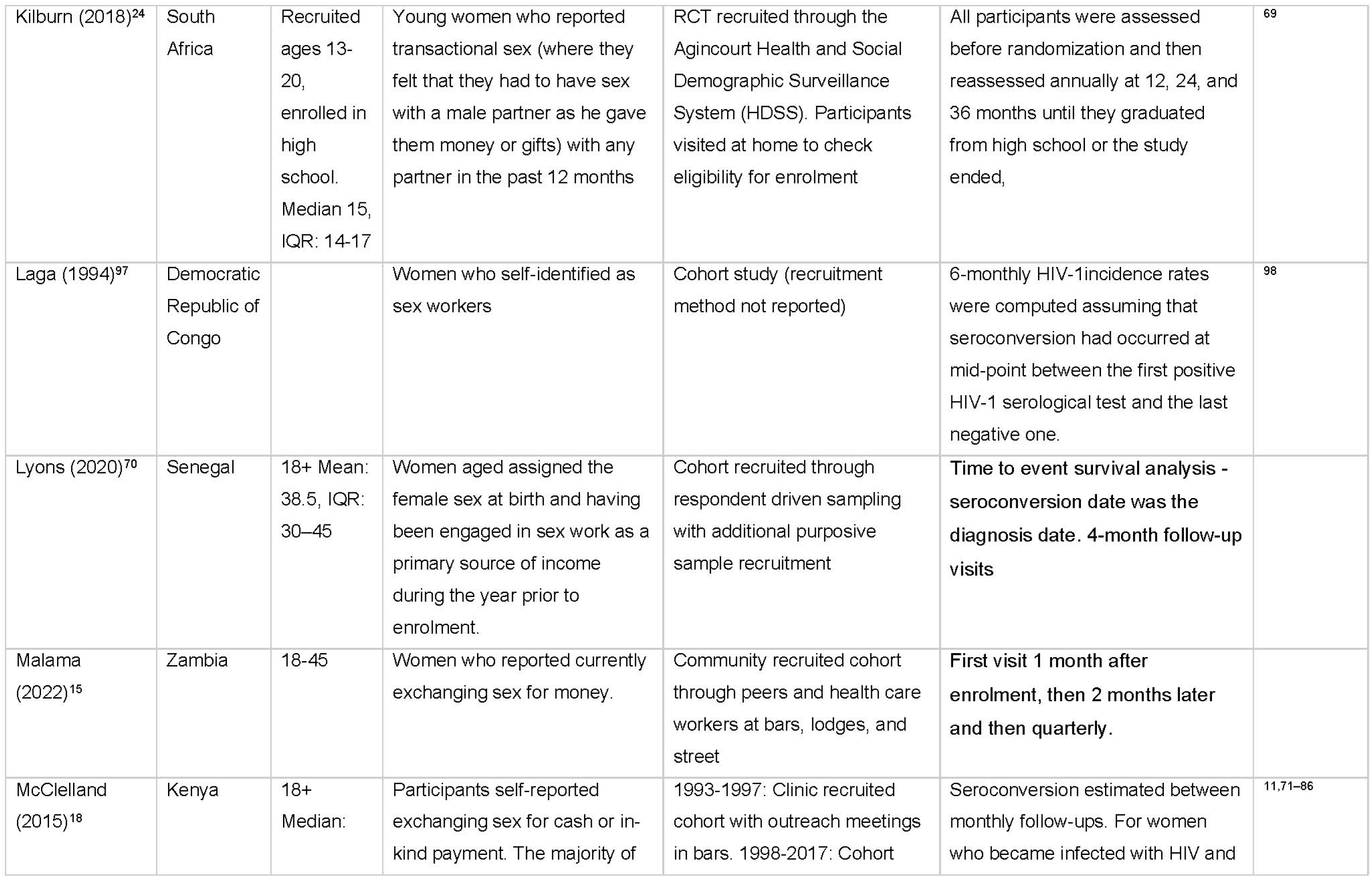
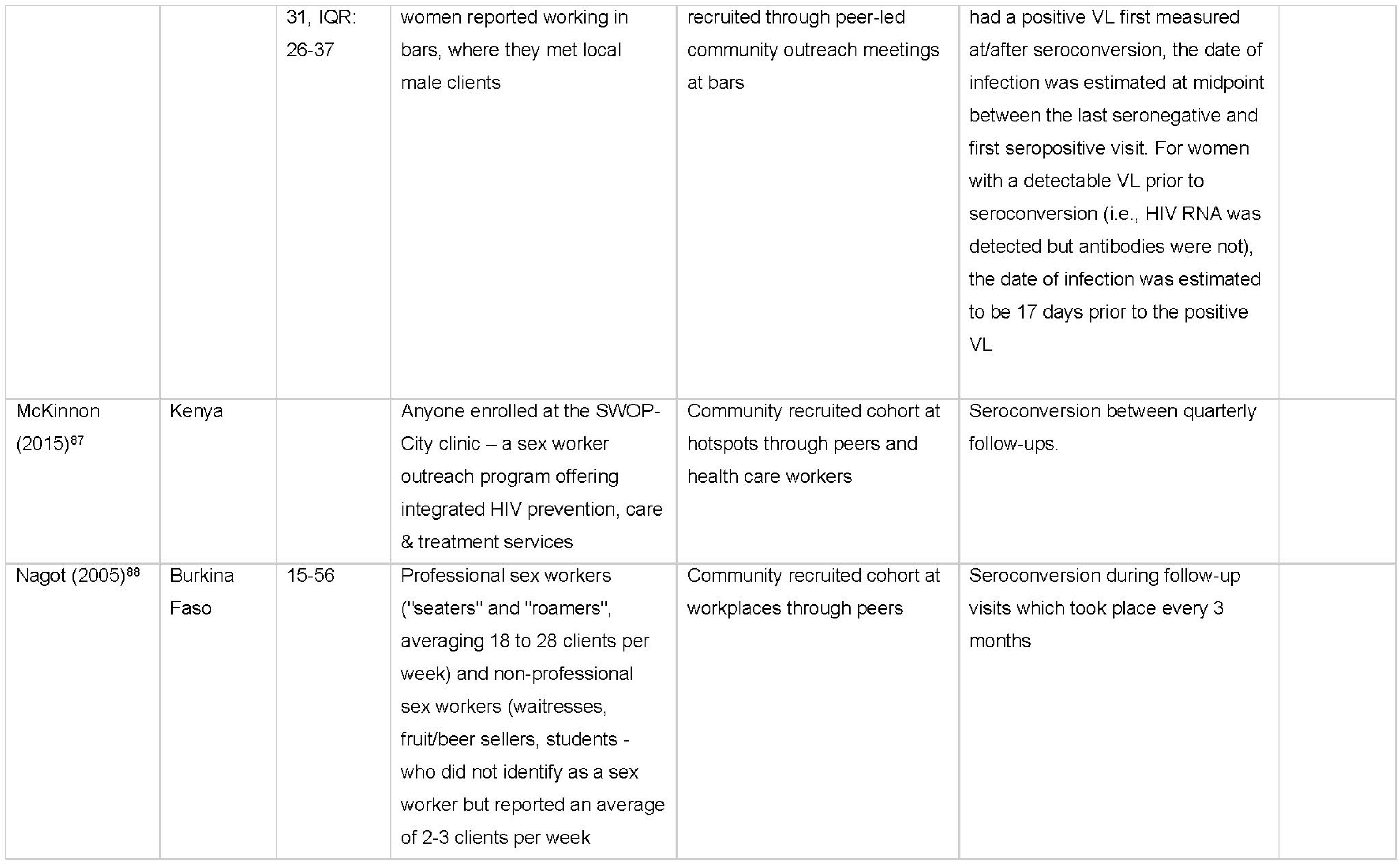
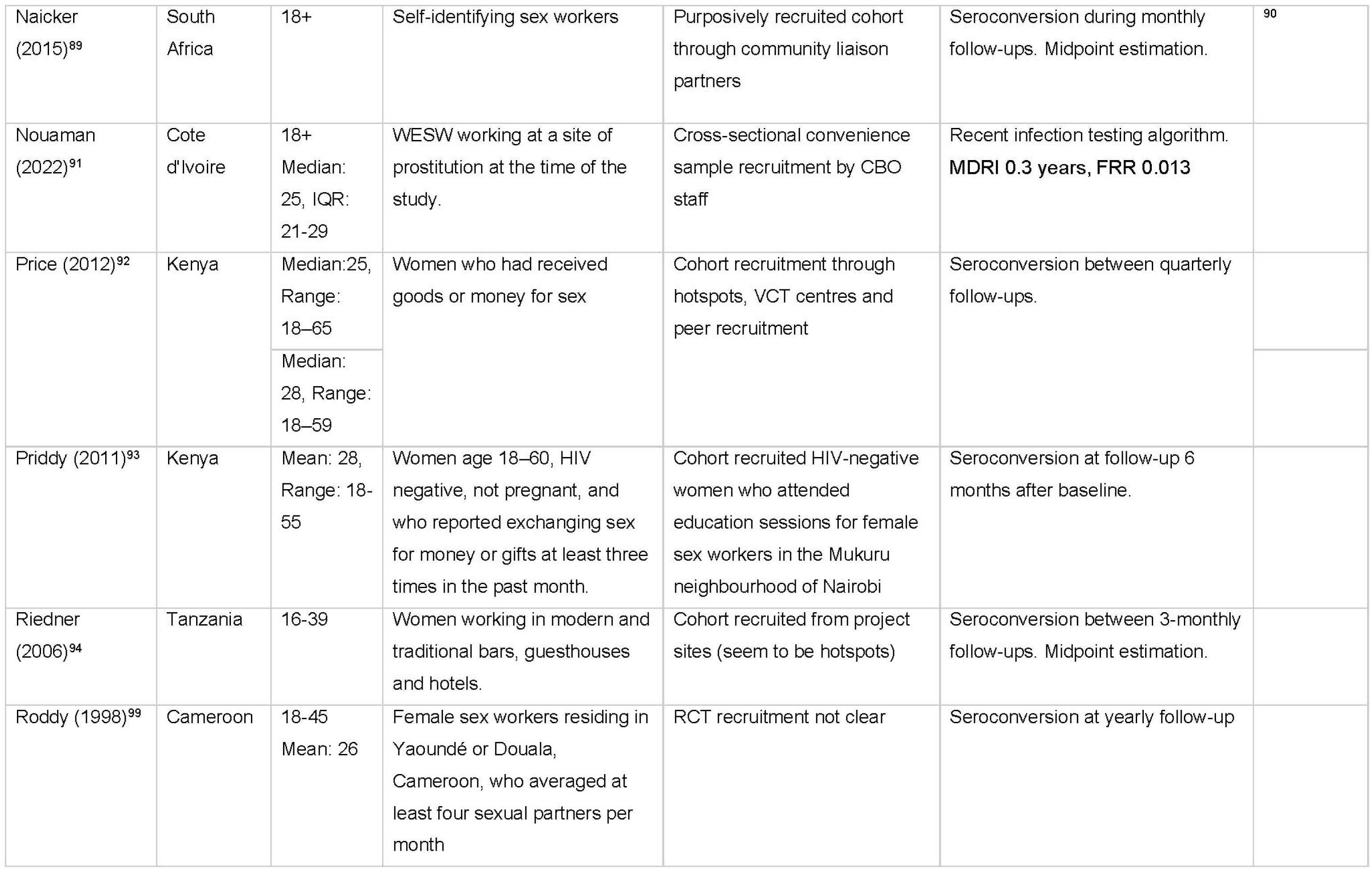
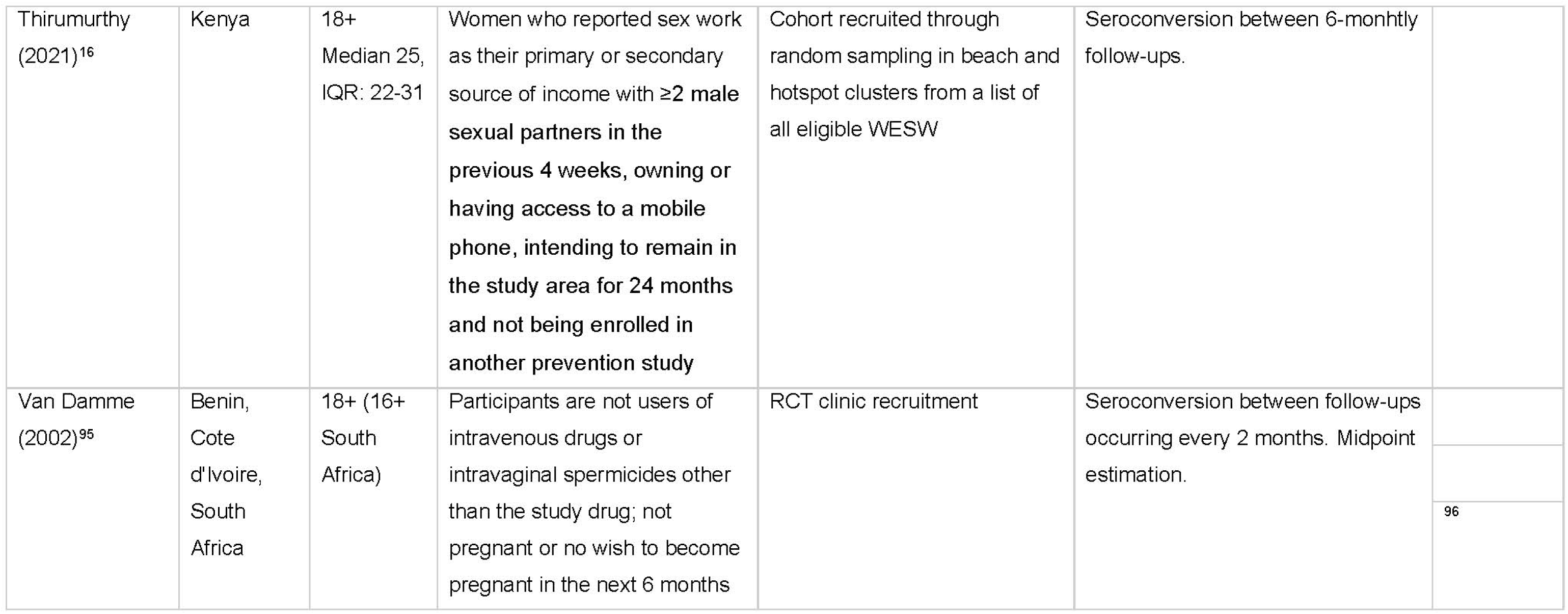
Study characteristics.

Over half the studies were cohort studies (59%; 19/32), one fifth were randomised control trials or intervention studies (22%; 7/32), five were cross-sectional studies (16%, 5/32), and one study used routine clinic data (1/32; Table 2). Study populations included women who self-identified as sex workers (7/32), exchanged sex for money or goods (13/32), either over a defined period or as a primary or secondary source of income, or worked in a known sex work location (2/32). Six study populations were women linked to clinics or sex worker programmes. Study reach varied widely, from studies in single clinic populations (6/32) to single towns, cities, or regions (19/32), or multiple locations nationally in South Africa and Zimbabwe (8/32). Recruitment methods included network sampling approaches (7/32), time location sampling (4/32), clinic recruitment (7/32), or convenience samples from peer outreach activities or community meetings (9/32). Study participant ages ranged from a median of 15 (IQR 14-17)^28^ to a mean of 38^29^ with 11/32 studies reporting a mean or median age between 25 and 30. Incidence estimates were predominantly derived from inference of a seroconversion date between HIV tests (26/32). Most of these studies used the midpoint between first positive and last negative test result (13/25), two used the HIV-positive test date, and seven did not report a clear seroconversion date estimation approach. Four studies estimated incidence from recency assays, and two used back calculation from age-specific HIV prevalence.

### Quality assessment

Assessment with the GHQAT classified 8/32 (25%) studies as ‘good’ quality, and the remaining 24/32 (77%) as ‘fair’ (Supplementary Table S2). Studies all scored well in terms of study design, with study objectives and study populations clearly defined. Most studies either had robust approaches to sampling or did not seek to generalise their findings beyond their study population so were considered adequately representative (26/32). Power calculations were presented in one-third of studies (9/32). Scores under study implementation varied, with only 10/31 studies reporting participation rates, of which six reported over 85% participation. For 19/32 studies, it was likely that participants enrolled were representative of the source population. Scoring for measurement of HIV incidence was variable. Among longitudinal studies, 12 report >70% participant retention at 12 months or study endpoint. Only five described methods to address loss to follow up.

### Data synthesis & meta-analysis

Incidence among WESW was correlated with matched female incidence (R = 0.41; Figure 2A). In meta-analysis, the HIV incidence rate in WESW was nine times higher than in matched total population women (RR 8.6, 95%CI 5.7–12.9; Figure 1). Rate ratios were greater in WCA (RR 22.4, 95%CI 11.3-44.3) than in ESA (RR 5.3, 95%CI 3.7–7.6). The sensitivity analysis using the 21 studies scoring above 60% through quality assessment yielded a pooled IRR of 6.9 (95% CI 4.4-10.8, Supplementary Figure 1). Sensitivity analysis using nationally-matched incidence increased relative risk (RR: 10.6, 95% CI 7.4-15.2, Supplementary Figure 2).

**Figure 2:**
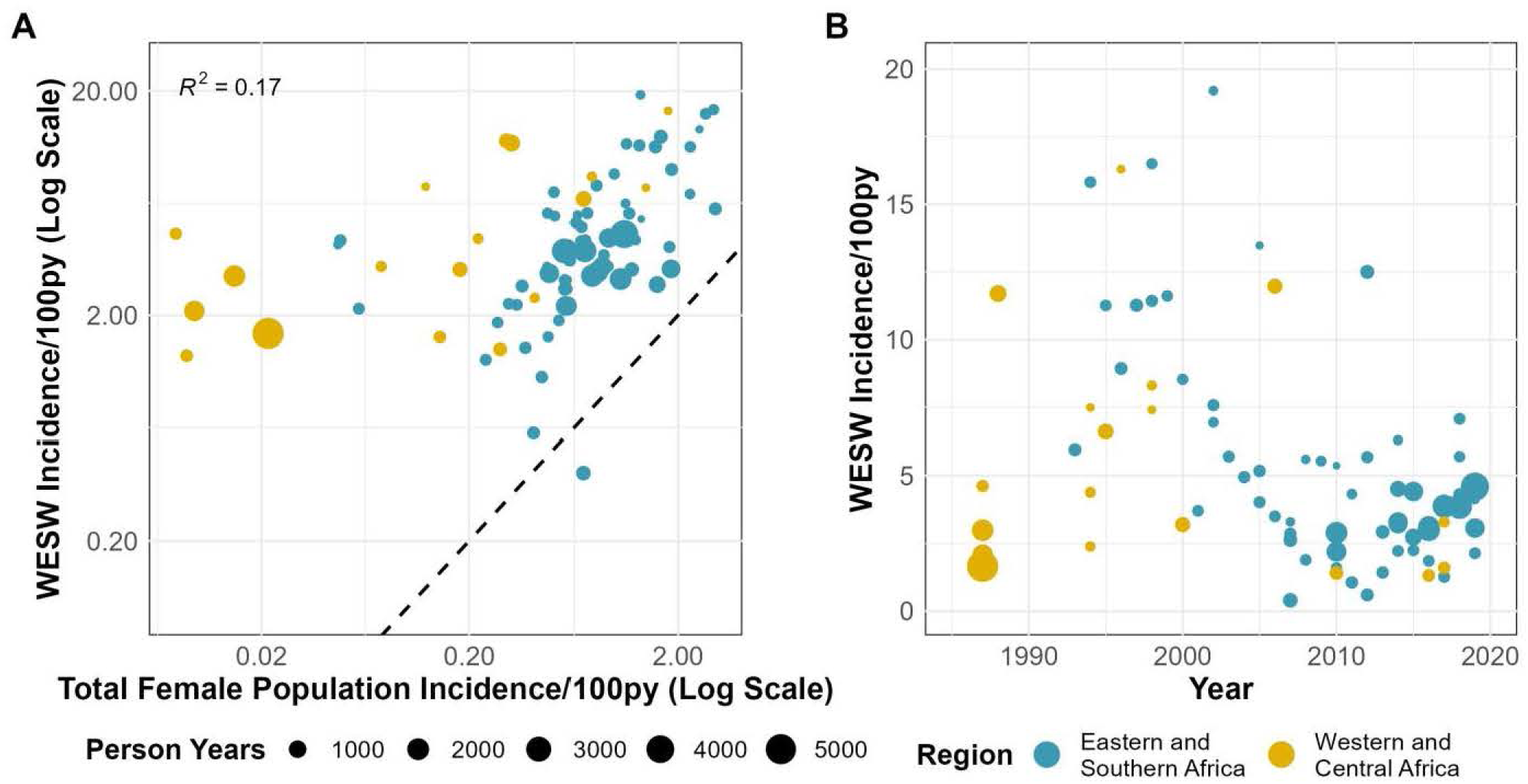
Empirical estimates of HIV Incidence among women engaging in sex work. A) Association between HIV incidence in sex workers and in the district-year-sex matched total population. B) Empirical estimates of HIV incidence in sex workers over time. Black dashed line in A represents the line of equality; WESW = women who engage in sex work; py = person-years

HIV incidence in WESW halved between 1985-1995 and 2010-2020, from a median of 5.96/100py (IQR 3.0-11.3/100py) to median 3.2/100py (IQR 2.2-4.3/100py; Figure 2B). Compared to matched female population incidence, there was no evidence for a change in IRR over time (Odds ratio [OR] 1.00, 95%CI 0.93-1.07; Figure 3). Incidence in WESW was nearly 6 times higher than in the total female population in 2003 according to the mixed-effects model (IRR 5.5, 95% CI 3.8-8.7; Figure 3, Supplementary Table S2).

**Figure 3.**
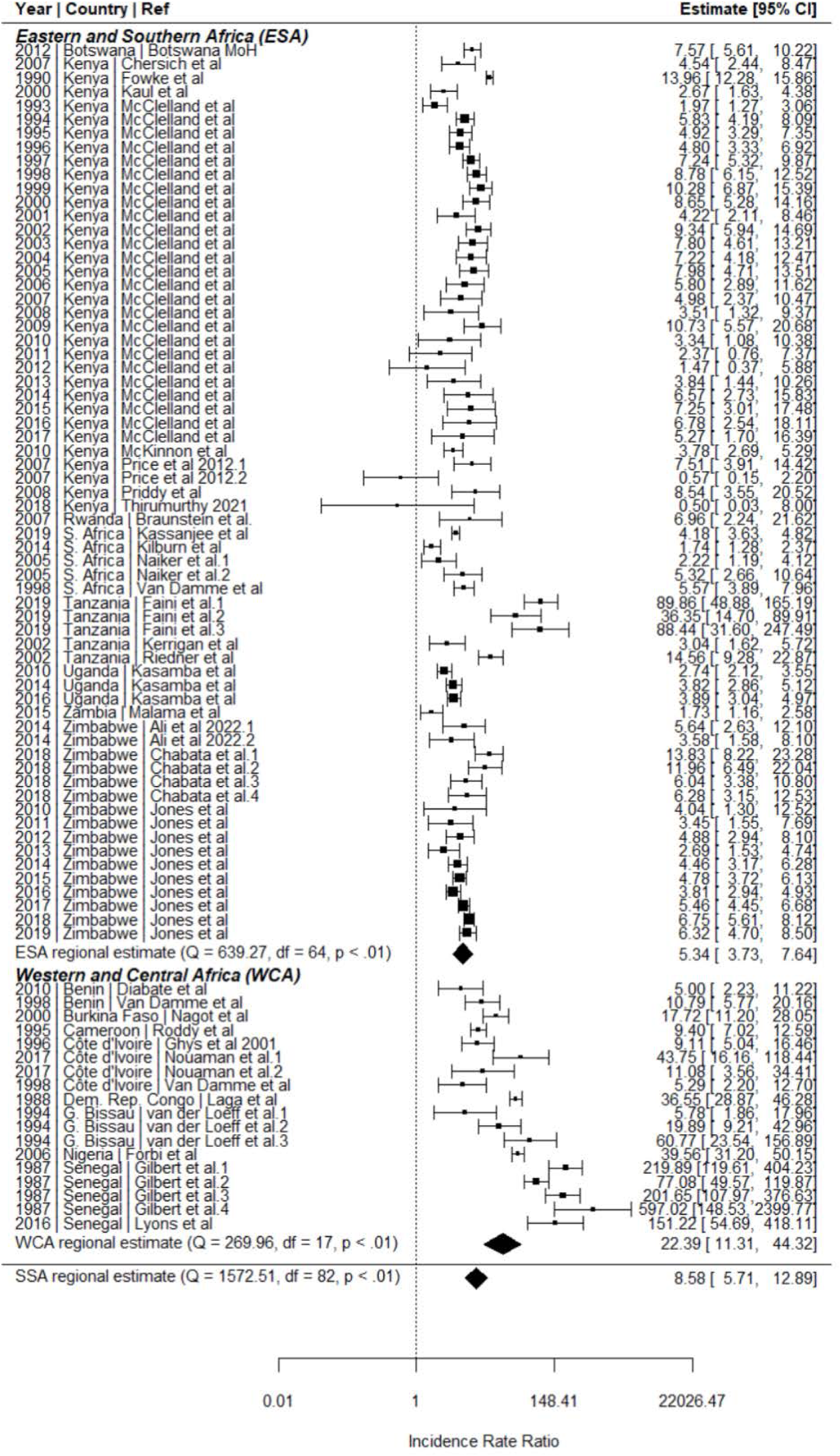
Meta-analysis of HIV incidence in women engaging in sex work relative to the total female population in Sub-Saharan Africa. IRRs calculated by dividing empirical estimates of sex worker HIV incidence by HIV incidence among age-district-year matched total population women derived from the district-level estimation model Naomi^21^. ESA: Eastern and Southern Africa; WCA: Western and Central Africa; SSA: Sub-Saharan Africa.

In separate model fits to data from Kenya and Zimbabwe, log IRR did not change over time in Kenya (OR 1.01, 95% CI: 1.00-1.03), but increased in Zimbabwe between 2009 and 2019 (OR 1.11, 95% CI 1.05-1.16; Supplementary Table S1). Two studies from these countries provided annual incidence estimates over their follow up-period (Figure 4). In the Kenyan cohort, incidence in WESW peaked at 16/100py in 1994 and declined to 1/100py in 2017. However, the declines in incidence in WESW were matched by reductions in total female population incidence, resulting in a stable IRR over the cohort period. In the Zimbabwean cohort, the HIV incidence rate in WESW also declined from 5/100py to 3/100py from 2010-2019, but the IRR increased due to faster relative declines in total population female incidence.

**Figure 4:**
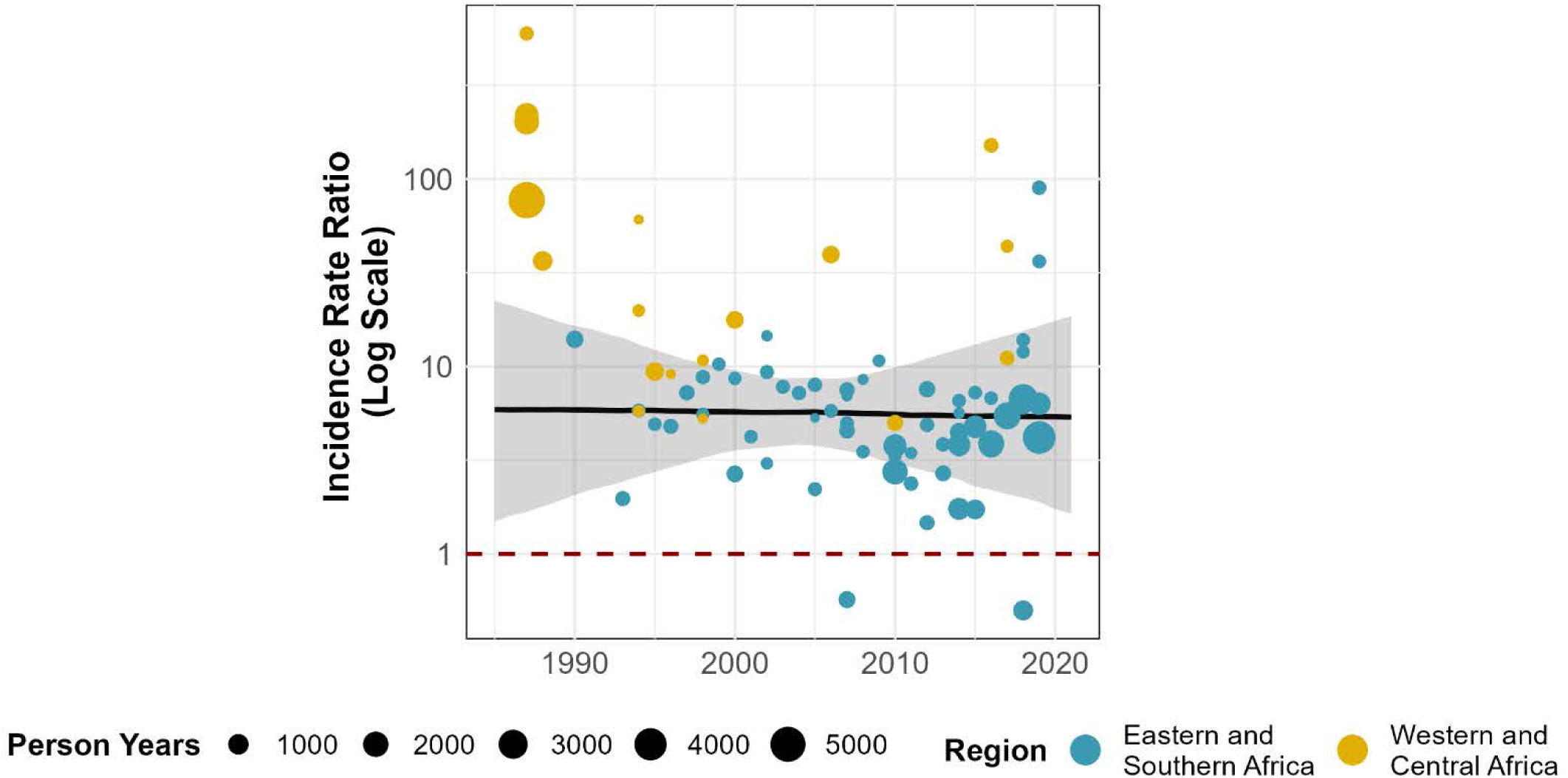
HIV incidence rate ratios modelled over time, presented on the logarithmic scale. Points represent incidence rate ratios calculated by dividing study-reported HIV incidence in women engaging in sex work by age-district-year matched total population HIV incidence derived from the Naomi model^21^. Black line represents the estimated incidence rate ratio (IRR) for Sub-Saharan Africa, with the grey shading capturing the 95% uncertainty range. The red dashed line represents an IRR of 1 (HIV incidence in sex workers = total female population HIV incidence).

**Figure 5:**
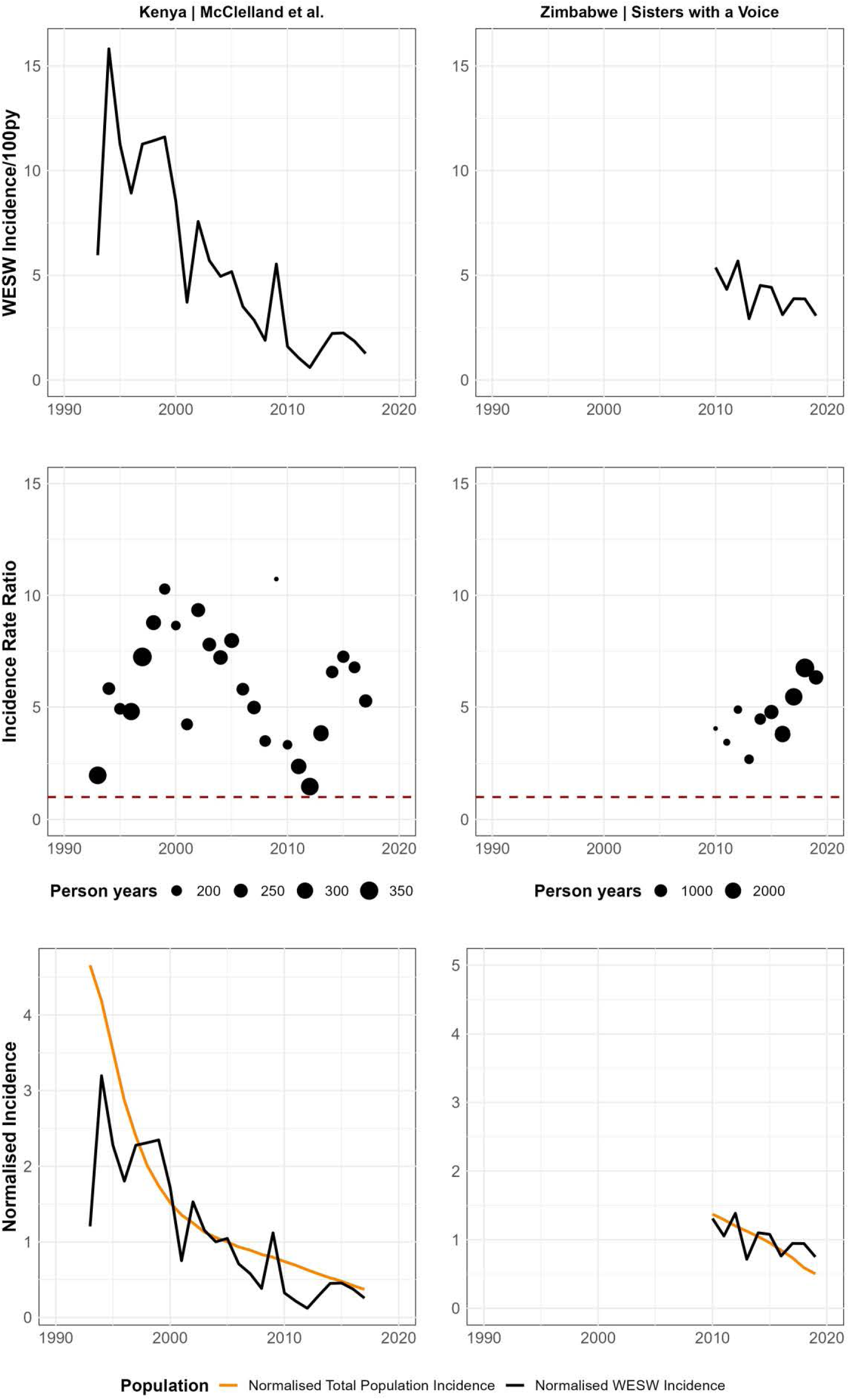
Case Studies - Incidence and Incidence Rate Ratio (IRR) trends over time in data from Mombasa, Kenya (McClelland et al) and Zimbabwe (Jones et al). Upper row: Empirical incidence in women engaging in sex work over time. Middle row: Incidence rate ratio over time. The red dashed line represents an IRR of 1 (HIV incidence in sex workers = total female population HIV incidence). Lower row: Normalised incidence over time. Calculated as a ratio of annual HIV incidence divided by incidence in the median year (1993-2018 for McClelland et al: median year = 2005; 2010-2019 for Jones et al: median year = 2015). WESW: Women who engage in sex work; IRR: Incidence Rate Ratio.

## Discussion

In sub-Saharan Africa, HIV incidence among women who engage in sex work was nearly nine times greater than in the wider female population. This disparity was larger in WCA, where incidence was almost 23 times higher than in all women, compared to six times higher in ESA. Between 1985 and 2020, incidence in WESW declined at a similar rate to incidence in age-location-year matched total population women, however case studies of Kenya and Zimbabwe illustrated that these trends may vary between countries. Sensitivity analyses using national incidence estimates and restricted to high-quality studies did not alter the interpretation of our findings.

Our analysis builds on existing evidence that WESW are disproportionately impacted by HIV. Our large IRR echoes the five-fold discrepancy reported in HIV prevalence between WESW and the wider population in sub-Saharan Africa.^2^ We additionally estimated a greater relative risk in lower HIV prevalence epidemics in WCA compared to ESA. The declining HIV incidence among WESW found in this analysis was aligned with modelling studies from WCA that estimate the proportion of total new HIV infections attributable to commercial sex has fallen over time.^30, 31^ These parallel incidence declines are in contrast with a systematic review of incidence among men who have sex with men in sub-Saharan Africa, which found no such evidence.^32^

Studies varied in their geographic reach, and their definition and recruitment of WESW. Inclusion criteria often defined the age range and duration or frequency of selling sex, or sought women self-identifying as WESW, leading to a range of study populations with likely varying degrees of HIV acquisition risk. Studies among women linked to targeted programmes or clinics may have recruited women at lower risk of HIV acquisition, who may be older or had sold sex for longer durations, potentially missing younger women, those yet to identify as selling sex and in the highest risk period immediately following sex work initiation.^33^ Preliminary findings from South Africa suggest that sex work dynamics are changing with the mean age of sex workers increasing and longer durations at-risk,^34^ whilst young WESW remain at highest incidence risk.^35^ Different sampling and recruitment approaches were also likely to identify different women for study inclusion. Studies have shown that participants recruited through RDS are younger than those recruited through venue-based snowball sampling.^36^ Using district-age-matched IRRs in our analysis likely mitigated some of the bias that would have come from geographical and age differences if raw estimates of incidence were analysed independently.

Whilst declining incidence in WESW may in part be a product of incidence trends and increased programme coverage across all women in SSA, it also reflects substantial efforts in roll-out of WESW-dedicated programmes, treatment, and health services in SSA, and rising treatment coverage among men.^1^ Whilst this finding is positive, our study also highlights the need to sustain and continue to expand effective HIV prevention options and treatment services for WESW, given the persistently large disparity in HIV incidence between WESW and the wider female population, particularly in WCA. Counterfactual-based modelling shows that fully meeting HIV prevention and treatment needs of WESW would substantially impact the HIV epidemic in SSA, and that averting HIV transmission associated with sex work should remain a programmatic priority.^6, 31^

Our study was subject to limitations. Data were available from fewer than half of SSA countries, with WCA particularly under-represented, and estimates for ESA largely from Kenya and Zimbabwe. This prevented estimation of country-specific IRRs and non-linear time trends and limited the generalisability of our findings. Data were insufficient to estimate pooled age-disaggregated IRRs which may have provided further insights, given evidence of higher incidence among younger WESW.^35^ For all but one study which provided a comparator non-WESW group,^24^ our IRR denominators were based on extrapolations of subnational estimates backwards in time parallel to national female incidence trajectories. Whilst this approach aimed to capture spatial variation over time, there is substantial uncertainty in these subnational incidence denominators, particularly for older studies, which is not reflected in the calculated IRRs. Additionally, given high mobility among WESW,^9^ matching on district boundaries may not yield the most comparable total female population denominator. However, sensitivity analysis using nationally-matched IRRs gave similar results to the district-matched analysis, alleviating concerns around subnational uncertainties.

We appraised studies using the GHQAT. Although we did not quantify changes in study quality over time, it is likely that study quality improved with improvements in HIV testing and sampling methods. Most studies did not make inferences beyond the study population they recruited and scored positively on adequately representing the wider WESW population. However, it is unlikely that many would have been representative had they made these inferences. Ascertaining the degree of bias associated with sample recruitment posed challenges. There is no standardised approach to sampling individuals from key populations, and although respondent driven sampling is widely accepted as a gold standard to achieving a representative sample,^38^ there was variation in implementation and reporting across studies. We excluded studies providing interventions beyond standard of care but were unable to account for wide variation in programme interventions and intensity, or the influence in historical or geographically proximal HIV responses.

Our review highlights that while HIV incidence data are available for WESW populations, geographical gaps remain, and temporal trends difficult to ascertain from WESW estimates alone, particularly at country level. Individual studies are challenging to compare due to substantial heterogeneity in study design and the limited generalisability of findings beyond individual study populations. Standardisation in definitions for WESW and reporting of age ranges may improve comparability of estimates. The application of methodological standards for directly measuring incidence in key populations would also be beneficial. Data gaps could be addressed by incorporating incidence measurements into survey and routine programmatic data analysis via serial cross-sectional prevalence data from biobehavioural surveys and recency testing,^12, 13, 17, 39^ instead of resource-intensive cohort studies. This would increase the number of available estimates to enable country-specific estimation and support real-time data-driven programming.

## Conclusion

Across SSA, HIV incidence in WESW has declined over the past 20 years but substantial inequalities remain, with incidence nine times greater among WESW compared to the wider female population. While declines are currently hard to ascertain from empirical estimates alone, these can be inferred from the limited change in WESW incidence relative to incidence among women in the wider population. Future surveillance activities should fill these gaps, ensuring a more standardised approach to obtaining empirical estimates of WESW incidence with repeat measures in the same populations to guide data-driven HIV prevention and treatment programme planning and ensure incidence continues to decline, and inequalities are addressed.

## Data Availability

All data produced in the present work are contained in the manuscript.

## Author contributions

OS, RLA, and HSJ conceptualised the study. JRH and HSJ conceived and conducted the initial literature review with HC. RLA updated the review and identified additional unpublished data. RLA led the quantitative data extraction and HSJ led data extraction of study methods. RSM, HT, FMC, and HSJ provided unpublished data. HSJ led the quality assessment. RLA and OS led the quantitative analysis. HSJ, RLA, and OS wrote the manuscript. JWI-E and JRH provided technical input throughout the study. All authors contributed to interpretation of results and edited the manuscript for intellectual content. All authors read and approved the final version of the manuscript for submission.

## Disclaimer

The findings here are those of the authors and do not necessarily represent the views or official position of the funding agencies.

## References

1. Joint United Nations Programme on HIV/AIDS (UNAIDS). The Path That Ends AIDS: UNAIDS Global AIDS Update 2023 | UNAIDS.; 2023. Accessed August 10, 2023. https://www.unaids.org/en/resources/documents/2023/global-aids-update-2023

2. Stevens O, Sabin K, Garcia SA, et al. Key population size, HIV prevalence, and ART coverage in sub-Saharan Africa: systematic collation and synthesis of survey data. medRxiv. Published online July 29, 2022:2022.07.27.22278071. doi:10.1101/2022.07.27.22278071

3. Beyrer C, Crago AL, Bekker LG, et al. An action agenda for HIV and sex workers. The Lancet. 2015;385(9964):287–301. doi:10.1016/S0140-6736(14)60933-8

4. Wheeler T, Wolf RC, Kapesa L, Cheng Surdo A, Dallabetta G. Scaling-Up HIV Responses with Key Populations in West Africa. JAIDS Journal of Acquired Immune Deficiency Syndromes. 2015;68(Supplement 2):S69–S73. doi:10.1097/QAI.0000000000000534

5. Cowan FM, Chabata ST, Musemburi S, et al. Strengthening the scale-up and uptake of effective interventions for sex workers for population impact in Zimbabwe. J Int AIDS Soc. 2019;22(S4). doi:10.1002/JIA2.25320/FULL

6. Stone J, Bothma R, Gomez GB, et al. Impact and cost-effectiveness of the national scale-up of HIV pre-exposure prophylaxis among female sex workers in South Africa: a modelling analysis. J Int AIDS Soc. 2023;26(2). doi:10.1002/JIA2.26063/FULL

7. Platt L, Grenfell P, Meiksin R, et al. Associations between sex work laws and sex workers’ health: A systematic review and meta-analysis of quantitative and qualitative studies. PLoS Med. 2018;15(12):e1002680. doi:10.1371/JOURNAL.PMED.1002680

8. UNAIDS. UNAIDS Guidance Note on HIV and Sex Work (Updated April 2012).; 2009. Accessed October 14, 2023. https://www.unaids.org/en/resources/documents/2012/20120402_UNAIDS-guidance-note-HIV-sex-work

9. Davey C, Dirawo J, Mushati P, Magutshwa S, Hargreaves JR, Cowan FM. Mobility and sex work: why, where, when? A typology of female-sex-worker mobility in Zimbabwe. Soc Sci Med. 2019;220:322–330. doi:10.1016/j.socscimed.2018.11.027

10. Malama K, Sagaon-Teyssier L, Gosset A, et al. Loss to follow-up among female sex workers in Zambia: findings from a five-year HIV-incidence cohort. African Journal of AIDS Research. 2020;19(4):296–303. doi:10.2989/16085906.2020.1836005

11. Graham SM, Raboud J, McClelland RS, et al. Loss to follow-up as a competing risk in an observational study of HIV-1 incidence. PLoS One. 2013;8(3). doi:10.1371/JOURNAL.PONE.0059480

12. Ali MS, Wit MDE, Chabata ST, et al. Estimation of HIV incidence from analysis of HIV prevalence patterns among female sex workers in Zimbabwe. AIDS. 2022;36(8):1141–1150. doi:10.1097/QAD.0000000000003198

13. Kassanjee R, Welte A, Otwombe K, et al. HIV incidence estimation among female sex workers in South Africa: a multiple methods analysis of cross-sectional survey data. Lancet HIV. 2022;9(11). doi:10.1016/S2352-3018(22)00201-6

14. Jones H, Cust H, Cowan F, Hensen B, Hargreaves J. Trends in HIV testing and knowledge of HIV status among female sex workers in sub-Saharan Africa: a systematic review. PROSPERO. Published 2020. Accessed October 14, 2023. https://www.crd.york.ac.uk/prospero/display_record.php?RecordID=162769

15. Malama K, Price MA, Sagaon-Teyssier L, et al. Evolution of Condom Use Among a 5-Year Cohort of Female Sex Workers in Zambia. AIDS Behav. 2022;26(2):470–477. doi:10.1007/S10461-021-03403-9

16. Thirumurthy H, Bair EF, Ochwal P, et al. The effect of providing women sustained access to HIV self-tests on male partner testing, couples testing, and HIV incidence in Kenya: a cluster-randomised trial. Lancet HIV. 2021;8(12):e736–e746. doi:10.1016/S2352-3018(21)00248-4

17. Jones HS, Hensen B, Musemburi S, et al. Interpreting declines in HIV test positivity: an analysis of routine data from Zimbabwe’s national sex work programme, 2009-2019. J Int AIDS Soc. 2022;25(7). doi:10.1002/JIA2.25943

18. McClelland RS, Richardson BA, Cherutich P, et al. A 15-year study of the impact of community antiretroviral therapy coverage on HIV incidence in Kenyan female sex workers. AIDS. 2015;29(17):2279–2286. doi:10.1097/QAD.0000000000000829

19. Rao A, Schwartz S, Viswasam N, et al. Evaluating the quality of HIV epidemiologic evidence for populations in the absence of a reliable sampling frame: a modified quality assessment tool. Ann Epidemiol. 2022;65:78. doi:10.1016/J.ANNEPIDEM.2021.07.009

20. UNAIDS, FHI 360, WHO, CDC, PEPFAR. Biobehavioural Survey Guidelines For Populations At Risk For HIV.; 2017. Accessed February 17, 2022. http://apps.who.int/bookorders.

21. UNAIDS. HIV sub-national estimates viewer. 2021. Accessed August 23, 2023. https://naomi-spectrum.unaids.org/

22. Stover J, Glaubius R, Teng Y, et al. Modeling the epidemiological impact of the UNAIDS 2025 targets to end AIDS as a public health threat by 2030. PLoS Med. 2021;18(10):e1003831. doi:10.1371/journal.pmed.1003831

23. UNAIDS. Spectrum file request – UNAIDS HIV Tools. Published 2023. Accessed August 24, 2023. https://hivtools.unaids.org/spectrum-file-request/

24. Kilburn K, Ranganathan M, Stoner MCD, et al. Transactional sex and incident HIV infection in a cohort of young women from rural South Africa. AIDS. 2018;32(12):1669–1677. doi:10.1097/QAD.0000000000001866

25. R: The R Project for Statistical Computing. Accessed February 7, 2023. https://www.r-project.org/

26. Viechtbauer W. The metafor Package: A Meta-Analysis Package for R. Accessed February 6, 2023. https://www.metafor-project.org/doku.php/metafor

27. R-INLA Project - What is INLA? Accessed February 6, 2023. https://www.r-inla.org/what-is-inla

28. Kilburn K, Ranganathan M, Stoner MCD, et al. Transactional sex and incident HIV infection in a cohort of young women from rural South Africa. AIDS. 2018;32(12):1669–1677. doi:10.1097/QAD.0000000000001866

29. Lyons CE, Olawore O, Turpin G, et al. Intersectional stigmas and HIV-related outcomes among a cohort of key populations enrolled in stigma mitigation interventions in Senegal. AIDS. 2020;34 Suppl 1(Suppl 1):S63–S71. doi:10.1097/QAD.0000000000002641

30. Mukandavire C, Walker J, Schwartz S, et al. Estimating the contribution of key populations towards the spread of HIV in Dakar, Senegal. J Int AIDS Soc. 2018;21(S5). doi:10.1002/jia2.25126

31. Maheu-Giroux M, Vesga JF, Diabaté S, et al. Changing Dynamics of HIV Transmission in Côte d’Ivoire: Modeling Who Acquired and Transmitted Infections and Estimating the Impact of Past HIV Interventions (1976–2015). JAIDS Journal of Acquired Immune Deficiency Syndromes. 2017;75(5):517–527. doi:10.1097/QAI.0000000000001434

32. Stannah J, Soni N, Keng J, et al. Trends in HIV testing, the treatment cascade, and HIV incidence among men who have sex with men in Africa: A systematic review and meta-regression analysis. medRxiv. Published online November 15, 2022:2022.11.14.22282329. doi:10.1101/2022.11.14.22282329

33. Neufeld B, Cholette F, Sandstrom P, et al. HIV acquisition prior to entry into formal sex work: inference from next-generation viral sequencing. AIDS. 2023;37(6):987. doi:10.1097/QAD.0000000000003484

34. Anderegg N, Johnson L. Increasing age and duration of sex work among female sex workers in South Africa and its potential impact: a meta-analysis and simulation exercise. IAS 2023. Published online 2023.

35. Stoner MCD, Rucinski KB, Lyons C, Napierala S. Differentiating the incidence and burden of HIV by age among women who sell sex: a systematic review and meta-analysis. J Int AIDS Soc. 2022;25(10). doi:10.1002/jia2.26028

36. Rao A, Stahlman S, Hargreaves J, et al. Sampling Key Populations for HIV Surveillance: Results From Eight Cross-Sectional Studies Using Respondent-Driven Sampling and Venue-Based Snowball Sampling. JMIR Public Health Surveill. 2017;3(4). doi:10.2196/PUBLICHEALTH.8116

37. Scorgie F, Chersich MF, Ntaganira I, Gerbase A, Lule F, Lo YR. Socio-Demographic Characteristics and Behavioral Risk Factors of Female Sex Workers in Sub-Saharan Africa: A Systematic Review. AIDS Behav. 2012;16(4):920–933. doi:10.1007/s10461-011-9985-z

38. Sabin K, Johnston LG, Sabin K. Sampling Hard-to-Reach Populations with Respondent Driven Sampling. Methodological Innovations Online. 2010;5(2):38.1–48. doi:10.4256/MIO.2010.0017

39. Botswana Ministry of Health. 2012 Mapping, Size Estimation & Behavioral and Biological Surveillance Survey (BBSS) of HIV/STI Among Select High-Risk Sub-Populations in Botswana BIOLOGICAL SURVEILLANCE SURVEY (BBSS) OF HIV/STI AMONG SELECT HIGH-RISK SUB-POPULATIONS IN BOTSWANA Technical Report July 2013 2 | P a g e RESULTS FROM THE 2012 BIOLOGICAL AND BEHAVIOURAL SURVEILLANCE SURVEY (BBSS) AMONG MOST AT RISK POPULATIONS IN BOTSWANA. Accessed November 22, 2022. https://www.fhi360.org/sites/default/files/media/documents/BBSS%202012%20Final.pdf

40. Braunstein SL, Ingabire CM, Kestelyn E, et al. High human immunodeficiency virus incidence in a cohort of Rwandan female sex workers. Sex Transm Dis. 2011;38(5):385–394. doi:10.1097/OLQ.0B013E31820B8EBA

41. Braunstein SL, van de Wijgert JH, Vyankandondera J, Kestelyn E, Ntirushwa J, Nash D. Risk Factor Detection as a Metric of STARHS Performance for HIV Incidence Surveillance Among Female Sex Workers in Kigali, Rwanda. Open AIDS J. 2012;6(1):112–121. doi:10.2174/1874613601206010112

42. Braunstein SL, Nash D, Kim AA, et al. Dual Testing Algorithm of BED-CEIA and AxSYM Avidity Index Assays Performs Best in Identifying Recent HIV Infection in a Sample of Rwandan Sex Workers. PLoS One. 2011;6(4). doi:10.1371/JOURNAL.PONE.0018402

43. Chabata ST, Hensen B, Chiyaka T, et al. The impact of the DREAMS partnership on HIV incidence among young women who sell sex in two Zimbabwean cities: results of a non-randomised study. BMJ Glob Health. 2021;6:3892. doi:10.1136/bmjgh-2020-003892

44. Chersich MF, Bosire W, King’ola N, Temmerman M, Luchters S. Effects of hazardous and harmful alcohol use on HIV incidence and sexual behaviour: a cohort study of Kenyan female sex workers. Global Health. 2014;10(1). doi:10.1186/1744-8603-10-22

45. Diabaté S, Chamberland A, Geraldo N, Tremblay C, Alary M. Gonorrhea, Chlamydia and HIV incidence among female sex workers in Cotonou, Benin: A longitudinal study. PLoS One. 2018;13(5). doi:10.1371/JOURNAL.PONE.0197251

46. Faini D, Msafiri F, Munseri P, et al. The Prevalence, Incidence, and Risk Factors for HIV Among Female Sex Workers-A Cohort Being Prepared for a Phase IIb HIV Vaccine Trial in Dar es Salaam, Tanzania. J Acquir Immune Defic Syndr (1988). 2022;91(5):439–448. doi:10.1097/QAI.0000000000003097

47. Forbi JC, Entonu PE, Mwangi LO, Agwale SM. Estimates of human immunodeficiency virus incidence among female sex workers in north central Nigeria: Implications for HIV clinical trials. Trans R Soc Trop Med Hyg. 2011;105(11):655–660. doi:10.1016/J.TRSTMH.2011.08.001/2/105-11-655-FIG002.GIF

48. Fowke KR, Nagelkerke NJD, Kimani J, et al. Resistance to HIV-1 infection among persistently seronegative prostitutes in Nairobi, Kenya. Lancet. 1996;348(9038):1347–1351. doi:10.1016/S0140-6736(95)12269-2

49. Ghys PD, Diallo MO, Ettiègne-Traoré V, et al. Effect of interventions to control sexually transmitted disease on the incidence of HIV infection in female sex workers. AIDS. 2001;15(11):1421–1431. doi:10.1097/00002030-200107270-00012

50. Gilbert PB, Mckeague IW, Eisen G, et al. Comparison of HIV-1 and HIV-2 infectivity from a prospective cohort study in Senegal. STATISTICS IN MEDICINE Statist Med. 2003;22:573–593. doi:10.1002/sim.1342

51. Kanki PJ, Travers KU, Marlink RG, et al. Slower heterosexual spread of HIV-2 than HIV-1. The Lancet. 1994;343(8903):943–946. doi:10.1016/S0140-6736(94)90065-5

52. Kanki PJ, Hamel DJ, Sankalé JL, et al. Human immunodeficiency virus type 1 subtypes differ in disease progression. J Infect Dis. 1999;179(1):68–73. doi:10.1086/314557

53. Travers K, Mboup S, Marlink R, et al. Natural Protection Against HIV-1 Infection Provided by HIV-2. Science (1979). 1995;268(5217):1612–1615. doi:10.1126/SCIENCE.7539936

54. Hargreaves JR, Mtetwa S, Davey C, et al. Cohort Analysis of Program Data to Estimate HIV Incidence and Uptake of HIV-Related Services Among Female Sex Workers in Zimbabwe, 2009-2014. J Acquir Immune Defic Syndr. 2016;72(1):e1–e8. doi:10.1097/QAI.0000000000000920

55. Kasamba I, Nash S, Shahmanesh M, et al. Missed Study Visits and Subsequent HIV Incidence Among Women in a Predominantly Sex Worker Cohort Attending a Dedicated Clinic Service in Kampala, Uganda. J Acquir Immune Defic Syndr. 2019;82(4):343–354. doi:10.1097/QAI.0000000000002143

56. Kasamba I, Nash S, Seeley J, Weiss HA. Human Immunodeficiency Virus Incidence Among Women at High-Risk of Human Immunodeficiency Virus Infection Attending a Dedicated Clinic in Kampala, Uganda: 2008-2017. Sex Transm Dis. 2019;46(6):407–415. doi:10.1097/OLQ.0000000000000978

57. Abaasa A, Mayanja Y, Asiki G, et al. Use of propensity score matching to create counterfactual group to assess potential HIV prevention interventions. Sci Rep. 2021;11(1). doi:10.1038/S41598-021-86539-X

58. Abaasa A, Nash S, Mayanja Y, et al. Simulated vaccine efficacy trials to estimate HIV incidence for actual vaccine clinical trials in key populations in Uganda. Published online 2019. doi:10.1016/j.vaccine.2019.02.072

59. Vandepitte J, Weiss HA, Bukenya J, et al. Association between Mycoplasma genitalium infection and HIV acquisition among female sex workers in Uganda: evidence from a nested case–control study. Sex Transm Infect. 2014;90(7):545. doi:10.1136/SEXTRANS-2013-051467

60. Redd AD, Ssemwanga D, Vandepitte J, et al. Rates of HIV-1 superinfection and primary HIV-1 infection are similar in female sex workers in Uganda. AIDS. 2014;28(14):2147–2152. doi:10.1097/QAD.0000000000000365

61. Kaul R, Kimani J, Nagelkerke NJ, et al. Monthly Antibiotic Chemoprophylaxis and Incidence of Sexually Transmitted Infections and HIV-1 Infection in Kenyan Sex Workers: A Randomized Controlled Trial. JAMA. 2004;291(21):2555–2562. doi:10.1001/JAMA.291.21.2555

62. Kaul R, Rutherford J, Rowland-Junes SL, et al. HIV-1 Env-specific cytotoxic T-lymphocyte responses in exposed, uninfected Kenyan sex workers: a prospective analysis. AIDS. 2004;18(15):2087–2089. doi:10.1097/00002030-200410210-00015

63. Peterson TA, Kimani J, Wachihi C, et al. HLA class I associations with rates of HIV-1 seroconversion and disease progression in the Pumwani Sex Worker Cohort. Tissue Antigens. 2013;81(2):93–107. doi:10.1111/TAN.12051

64. Plummer FA, Chubb H, Simonsen JN, et al. Antibodies to opacity proteins (Opa) correlate with a reduced risk of gonococcal salpingitis. J Clin Invest. 1994;93(4):1748–1755. doi:10.1172/JCI117159

65. Plummer FA, Simonsen JN, Cameron DW, et al. Cofactors in Male-Female Sexual Transmission of Human Immunodeficiency Virus Type 1. J Infect Dis. 1991;163(2):233–239. doi:10.1093/INFDIS/163.2.233

66. Willerford DM, Bwayo JJ, Hensel M, et al. Human immunodeficiency virus infection among high-risk seronegative prostitutes in Nairobi. J Infect Dis. 1993;167(6):1414–1417. doi:10.1093/INFDIS/167.6.1414

67. MacDonald KS, Fowke KR, Kimani J, et al. Influence of HLA supertypes on susceptibility and resistance to human immunodeficiency virus type 1 infection. J Infect Dis. 2000;181(5):1581–1589. doi:10.1086/315472

68. Kerrigan D, Mbwambo J, Likindikoki S, et al. Project Shikamana: Community Empowerment-Based Combination HIV Prevention Significantly Impacts HIV Incidence and Care Continuum Outcomes Among Female Sex Workers in Iringa, Tanzania. J Acquir Immune Defic Syndr. 2019;82(2):141–148. doi:10.1097/QAI.0000000000002123

69. Ranganathan M, Kilburn K, Stoner MCD, et al. The Mediating Role of Partner Selection in the Association Between Transactional Sex and HIV Incidence Among Young Women. J Acquir Immune Defic Syndr. 2020;83(2):103–110. doi:10.1097/QAI.0000000000002225

70. Laga M, Alary M, Behets F, et al. Condom promotion, sexually transmitted diseases treatment, and declining incidence of HIV-1 infection in female Zairian sex workers. Lancet. 1994;344(8917):246–248. doi:10.1016/S0140-6736(94)93005-8

71. Laga M, Manoka A, Kivuvu M, et al. Non-ulcerative sexually transmitted diseases as risk factors for HIV-1 transmission in women: results from a cohort study. AIDS. 1993;7(1):95–102. doi:10.1097/00002030-199301000-00015

72. Lyons CE, Olawore O, Turpin G, et al. Intersectional stigmas and HIV-related outcomes among a cohort of key populations enrolled in stigma mitigation interventions in Senegal. AIDS. 2020;34(Suppl 1):S63. doi:10.1097/QAD.0000000000002641

73. Baeten JM, Benki S, Chohan V, et al. Hormonal contraceptive use, herpes simplex virus infection, and risk of HIV-1 acquisition among Kenyan women. AIDS. 2007;21(13):1771–1777. doi:10.1097/QAD.0B013E328270388A

74. Baeten JM, Lavreys L, Sagar M, et al. Effect of contraceptive methods on natural history of HIV: studies from the Mombasa cohort. J Acquir Immune Defic Syndr. 2005;38 Suppl 1. doi:10.1097/01.QAI.0000167030.18278.0E

75. Graham SM, Raboud J, Jaoko W, Mandaliya K, McClelland RS, Bayoumi AM. Changes in Sexual Risk Behavior in the Mombasa Cohort: 1993–2007. PLoS One. 2014;9(11). doi:10.1371/JOURNAL.PONE.0113543

76. Lavreys L, Thompson M Lou, Martin HL, et al. Primary human immunodeficiency virus type 1 infection: clinical manifestations among women in Mombasa, Kenya. Clin Infect Dis. 2000;30(3):486–490. doi:10.1086/313718

77. Lavreys L, Baeten JM, Overbaugh J, et al. Virus load during primary human immunodeficiency virus (HIV) type 1 infection is related to the severity of acute HIV illness in Kenyan women. Clinical Infectious Diseases. 2002;35(1):77–81. doi:10.1086/340862/2/35-1-77-FIG001.GIF

78. Martin HL, Nyange PM, Richardson BA, et al. Hormonal Contraception, Sexually Transmitted Diseases, and Risk of Heterosexual Transmission of Human Immunodeficiency Virus Type 1. J Infect Dis. 1998;178(4):1053–1059. doi:10.1086/515654

79. Martin HL, Richardson BA, Mandaliya K, Achola JO, Overbaugh J, Kreiss JK. The early work on hormonal contraceptive use and HIV acquisition. J Acquir Immune Defic Syndr. 2005;38 Suppl 1(SUPPL. 1). doi:10.1097/01.QAI.0000167027.33525.1C

80. Martin HL, Richardson BA, Nyange PM, et al. Vaginal Lactobacilli, Microbial Flora, and Risk of Human Immunodeficiency Virus Type 1 and Sexually Transmitted Disease Acquisition. J Infect Dis. 1999;180(6):1863–1868. doi:10.1086/315127

81. McClelland RS, Hassan WM, Lavreys L, et al. HIV-1 acquisition and disease progression are associated with decreased high-risk sexual behaviour among Kenyan female sex workers. AIDS. 2006;20(15):1969–1973. doi:10.1097/01.AIDS.0000247119.12327.E6

82. McClelland RS, Lavreys L, Hassan WM, Mandaliya K, Ndinya-Achola JO, Baeten JM. Vaginal washing and increased risk of HIV-1 acquisition among African women: A 10-year prospective study. AIDS. 2006;20(2):269–273. doi:10.1097/01.AIDS.0000196165.48518.7B

83. McClelland RS, Lavreys L, Katingima C, et al. Contribution of HIV-1 infection to acquisition of sexually transmitted disease: A 10-year prospective study. Journal of Infectious Diseases. 2005;191(3):333–338. doi:10.1086/427262/2/191-3-333-TAB002.GIF

84. McClelland RS, Sangaré L, Hassan WM, et al. Infection with Trichomonas vaginalis increases the risk of HIV-1 acquisition. J Infect Dis. 2007;195(5):698–702. doi:10.1086/511278

85. McClelland RS, Lingappa JR, Srinivasan S, et al. Evaluation of the association between the concentrations of key vaginal bacteria and the increased risk of HIV acquisition in African women from five cohorts: a nested case-control study. Lancet Infect Dis. 2018;18(5):554–564. doi:10.1016/S1473-3099(18)30058-6

86. Richardson BA, Lavreys L, Martin HL, et al. Evaluation of a low-dose nonoxynol-9 gel for the prevention of sexually transmitted diseases: a randomized clinical trial. Sex Transm Dis. 2001;28(7):394–400. doi:10.1097/00007435-200107000-00006

87. Sabo MC, Richardson BA, Lavreys L, et al. Does bacterial vaginosis modify the effect of hormonal contraception on HIV seroconversion. AIDS. 2019;33(7):1225–1230. doi:10.1097/QAD.0000000000002167

88. Willcox AC, Richardson BA, Shafi J, et al. Derivation of an HIV Risk Score for African Women Who Engage in Sex Work. AIDS Behav. 2021;25(10):3292–3302. doi:10.1007/S10461-021-03235-7

89. McKinnon LR, Izulla P, Nagelkerke N, et al. Risk Factors for HIV Acquisition in a Prospective Nairobi-Based Female Sex Worker Cohort. AIDS Behav. 2015;19(12):2204–2213. doi:10.1007/S10461-015-1118-7

90. Nagot N, Ouedraogo A, Ouangre A, et al. Is sexually transmitted infection management among sex workers still able to mitigate the spread of HIV infection in West Africa? J Acquir Immune Defic Syndr (1988). 2005;39(4):454–458. doi:10.1097/01.QAI.0000152399.54648.B9

91. Naicker N, Kharsany ABM, Werner L, et al. Risk factors for HIV acquisition in high risk women in a generalised epidemic setting. AIDS Behav. 2015;19(7):1305. doi:10.1007/S10461-015-1002-5

92. van Loggerenberg F, Mlisana K, Williamson C, et al. Establishing a cohort at high risk of HIV infection in South Africa: challenges and experiences of the CAPRISA 002 acute infection study. PLoS One. 2008;3(4). doi:10.1371/JOURNAL.PONE.0001954

93. Nouaman MN, Becquet V, Plazy M, et al. Incidence of HIV infection and associated factors among female sex workers in Côte d’Ivoire, results of the ANRS 12361 PrEP-CI study using recent infection assays. PLoS One. 2022;17(11). doi:10.1371/JOURNAL.PONE.0271988

94. Price MA, Rida W, Mwangome M, et al. Identifying at-risk populations in kenya and south africa: HIV incidence in cohorts of menwho report sex with men, sex workers, and youth. J Acquir Immune Defic Syndr (1988). 2012;59(2):185–193. doi:10.1097/QAI.0B013E31823D8693

95. Priddy FH, Wakasiaka S, Hoang TD, et al. Anal Sex, Vaginal Practices, and HIV Incidence in Female Sex Workers in Urban Kenya: Implications for the Development of Intravaginal HIV Prevention Methods. AIDS Res Hum Retroviruses. 2011;27(10):1067. doi:10.1089/AID.2010.0362

96. Riedner G, Hoffmann O, Rusizoka M, et al. Decline in sexually transmitted infection prevalence and HIV incidence in female barworkers attending prevention and care services in Mbeya Region, Tanzania. AIDS. 2006;20(4):609–615. doi:10.1097/01.AIDS.0000210616.90954.47

97. Roddy RE, Zekeng L, Kelley RA, Tamoufé U, Weir SS, Wong EL. A Controlled Trial of Nonoxynol 9 Film to Reduce Male-to-Female Transmission of Sexually Transmitted Diseases. 101056/NEJM199808203390803. 1998;339(8):504–510. doi:10.1056/NEJM199808203390803

98. 98. Van Damme L, Ramjee G, Alary M, et al. Effectiveness of COL-1492, a nonoxynol-9 vaginal gel, on HIV-1 transmission in female sex workers: A randomised controlled trial. Lancet. 2002;360(9338):971–977. doi:10.1016/S0140-6736(02)11079-8

99. Auvert B, Marais D, Lissouba P, Zarca K, Ramjee G, Williamson AL. High-risk human papillomavirus is associated with HIV acquisition among South African female sex workers. Infect Dis Obstet Gynecol. 2011;2011. doi:10.1155/2011/692012

