## Supplementary File for "HIV incidence among women engaging in sex work in sub-Saharan Africa: a systematic review and meta-analysis"

**Supplementary text 1: Search strategies for the reviews.**

**Initial search:** 19 June 2019; Date restrictions applied: 1 January 1990 – 19 June 2019

| **MEDLINE Searches (Ovid MEDLINE(R) and Epub Ahead of Print, In-Process & Other Non-Indexed Citations, Daily and Versions(R) 1946 to June 04, 2019) (saved)** | | |
|---|---|---|
| **Sex Workers** | **HIV** | **sub-Saharan Africa** |
| 1. Sex Work/ | 1. exp HIV/ | 1. africa/ or exp "africa south of the sahara"/ |
| 2. Sex Workers/ | 2. Acquired Immunodeficiency Syndrome/ | 2. Africa.mp. |
| 3. (sex adj2 work*).mp. | 3. exp HIV Seroprevalence/ | 3. (((Angola or Benin or Botswana or Burkina Faso or Burundi or Cabo Verde or Cameroon or Central African Republic or Chad or Comoros or Dem* Rep* Congo or Rep* Congo or Cote D'ivoire or Equatorial Guinea or Eritrea or Eswatini or Ethiopia or Gabon or Gambia or Ghana or Guinea or Guinea-Bissau or Kenya or Lesotho or Liberia or Madagascar or Malawi or Mali or Mauritania or Mauritius or Mozambique or Namibia or Niger or Nigeria or Rwanda or Sao Tome) and Principe) or Senegal or Seychelles or Sierra Leone or Somalia or South Africa or South Sudan or Sudan or Tanzania or Togo or Uganda or Zambia or Zimbabwe).mp. |
| 4. FSW*.mp. | 4. exp HIV Seropositivity/ or exp HIV Infections/ | |
| 5. prostitut*.mp. | 5. HIV.mp. | |
| 6. transactional sex.mp. | 6. human immun* virus.mp. | |
| 7. commercial sex.mp. | 7. AIDS.mp. | |
| 8. sell* sex.mp. | 8. acquire* immun* syndrome.mp. | |
| 9. (paid adj3 sex).mp. | | |
| 10. sex industry.mp. | | |
| 11. key population*.mp. | | |
| 12. (high risk adj2 women).mp. | | |
| 13. (high risk adj2 girls).mp. | | |
| [mp=title, abstract, original title, name of substance word, subject heading word, floating sub-heading word, keyword heading word, organism supplementary concept word, protocol supplementary concept word, rare disease supplementary concept word, unique identifier, synonyms] | | |

| **EMBASE via Ovid** | | |
|---|---|---|
| **Sex Workers** | **HIV** | **sub-Saharan Africa** |
| 1. prostitution/ | 1. Human immunodeficiency virus/ | 1. Africa/ |
| 2. sex worker/ | 2. acquired immune deficiency syndrome/ | 2. exp "Africa south of the Sahara"/ |
| 3. (sex adj2 work*).mp. | 3. HIV.mp. | 3. Africa.mp. |
| 4. FSW*.mp. | 4. human immun* virus.mp. | 4. (((Angola or Benin or Botswana or Burkina Faso or Burundi or Cabo Verde or Cameroon or Central African Republic or Chad or Comoros or Dem* Rep* Congo or Rep* Congo or Cote D'ivoire or Equatorial Guinea or Eritrea or Eswatini or Ethiopia or Gabon or Gambia or Ghana or Guinea or Guinea- |
| 5. prostitut*.mp. | 5. AIDS.mp. | |
| 6. transactional sex.mp. | 6. acquire* immun* syndrome.mp. | |
| 7. commercial sex.mp. | | |
| 8. sell* sex.mp. | | |
| 9. (paid adj3 sex).mp. | | |
| 10. sex industry.mp. | | |

| | |
|---|---|
| 11. key population*.mp. | Bissau or Kenya or Lesotho or Liberia or Madagascar or Malawi or Mali or Mauritania or Mauritius or Mozambique or Namibia or Niger or Nigeria or Rwanda or Sao Tome) and Principe) or Senegal or Seychelles or Sierra Leone or Somalia or South Africa or South Sudan or Sudan or Tanzania or Togo or Uganda or Zambia or Zimbabwe).mp. [mp=title, abstract, heading word, drug trade name, original title, device manufacturer, drug manufacturer, device trade name, keyword, floating subheading word, candidate term word] |
| 12. (high risk adj2 women).mp. | |
| 13. (high risk adj2 girls).mp. | |

| **Web of Science** | | |
|---|---|---|
| **Sex Workers** | **HIV** | **sub-Saharan Africa** |
| (TS= (girls at high risk)) | (TS=(human immun* virus)) | (TS=Africa) AND LANGUAGE: (English) |
| (TS= (high risk girls)) | (TS=(acquire* immun* deficiency syndrome)) | |
| (TS= (women at high risk)) | (TS=AIDS) | |
| (TS= (high risk women)) | (TS=HIV) | |
| (TS=(key population*)) | (TS=acquire* immun* deficiency syndrome) | |
| (TS= (sex industry)) | (TS=human immun* virus) | |
| (TS= (paid for sex)) | | |
| (TS= (sell* sex)) | | |
| (TS=(commercial sex)) | | |
| (TS= (transactional sex)) | | |
| (TS=prostitut*) | | |
| (TS=FSW*) | | |
| (TS= (sex work*)) | | |
| Indexes=SCI-EXPANDED, SSCI, A&HCI, CPCI-S, CPCI-SSH, ESCI Timespan=1990-2019 DOCUMENT TYPES: (Article) | | |

| **Global Health via Ovid** | | |
|---|---|---|
| **Sex Workers** | **HIV** | **sub-Saharan Africa** |
| 1. exp prostitutes/ or prostitution/ | 1. human immunodeficiency viruses/ or hiv infections/ or human immunodeficiency virus 1/ or human immunodeficiency virus 2/ | 1. africa/ or exp "africa south of sahara"/ |
| 2. exp sex workers/ | 2. acquired immune deficiency syndrome/ | 2. Africa.mp. [mp=abstract, title, original title, broad terms, heading words, identifiers, cabicodes] |
| 3. (sex adj2 work*).mp. | 3. HIV.mp. | 3. (((Angola or Benin or Botswana or Burkina Faso or Burundi or Cabo Verde or Cameroon or Central African Republic or Chad or Comoros or Dem* Rep* Congo or Rep* Congo or Cote D'ivoire or Equatorial Guinea or Eritrea or Eswatini or Ethiopia or Gabon or Gambia or Ghana or Guinea or Guinea-Bissau or Kenya or Lesotho or Liberia or |
| 4. FSW*.mp. | 4. human immun* virus.mp. | |
| 5. prostitut*.mp. | 5. AIDS.mp. | |
| 6. transactional sex.mp. | 6. acquire* immun* syndrome.mp. | |
| 7. commercial sex.mp. | | |
| 8. sell* sex.mp. | | |
| 9. (paid adj3 sex).mp. | | |
| 10. sex industry.mp. | | |

| | |
|---|---|
| 11. key population*.mp. | Madagascar or Malawi or Mali or Mauritania or Mauritius or Mozambique or Namibia or Niger or Nigeria or Rwanda or Sao Tome) and Principe) or Senegal or Seychelles or Sierra Leone or Somalia or South Africa or South Sudan or Sudan or Tanzania or Togo or Uganda or Zambia or Zimbabwe).mp. [mp=abstract, title, original title, broad terms, heading words, identifiers, cabicodes] |
| 12. (high risk adj2 women).mp. | |
| 13. (high risk adj2 girls).mp. | |

**Updated search:** 3 January 2023; Date restrictions applied: 1 January 1990 – 31 December 2022

**Medline:**

1. (Kigali or Soweto or Johannesburg or Durban or Port Elizabeth or Mpumalanga or Abidjan or Harare or Mombasa or Nairobi or Brazzaville or “Dar es Salaam” or “Cape Town” or “Addis Ababa” or Douala or Accra or Luanda or Lusaka or Conakry or Kampala or Maputo or Freetown or Bangui or Bosaso).mp. [mp=abstract, title, original title, heading words, cabicodes words]
2. (Angola or Botswana or Eswatini or Swaziland or Ethiopia or Kenya or Lesotho).mp. [mp=abstract, title, original title, heading words, cabicodes words]
3. Malawi.mp.
4. (Mozambique or Namibia).mp. [mp=abstract, title, original title, heading words, cabicodes words]
5. “South Africa”.mp.
6. (“South Sudan” or Uganda or Tanzania).mp. [mp=abstract, title, original title, heading words, cabicodes words]
7. (Zambia or Zimbabwe or Benin or “Burkina Faso” or Burundi or Cameroon or “Central African Republic” or Chad or Congo or “Cote d’Ivoire” or “Ivory Coast” or “Democratic Republic of the Congo” or “Equatorial Guinea” or Gabon or Gambia or Ghana or Guinea or “Guinea-Bissau” or Liberia or Mali or Niger or Nigeria or Senegal or “Sierra Leone” or Togo).mp. [mp=abstract, title, original title, heading words, cabicodes words]
8. “Africa South of the Sahara”/
9. 1 or 2 or 3 or 4 or 5 or 6 or 7 or 8
10. exp HIV/
11. (HIV or “Human Immunodeficiency Virus” or “HIV-1”).mp. [mp=abstract, title, original title, heading words, cabicodes words]
12. (incidence or prospective or cohort or longitudinal or panel or rostitute* or sero-conver*).mp. [mp=abstract, title, original title, heading words, cabicodes words]
13. (“female sex workers” or “female sex worker” or “women who sell sex” or “woman who sells sex” or prostitute* or “sex-worker” or “sex work” or “sex-work” or FSW).mp. [mp=abstract, title, original title, heading words, cabicodes words]
14. 10 or 11 or 12
15. 9 and 13 and 14
16. limit 15 to yr=”1990 – 2022”

**Global Health:**

1. ((Kigali or Soweto or Johannesburg or Durban or Port Elizabeth or Mpumalanga or Abidjan or Harare or Mombasa or Nairobi or Brazzaville or "Dar es Salaam" or "Cape Town" or "Addis Ababa" or Douala or Accra or Luanda or Lusaka or Conakry or Kampala or Maputo or Freetown or Bangui or Bosaso or Angola or Botswana or Eswatini or Swaziland or Ethiopia or Kenya or Lesotho or Malawi or Mozambique or Namibia or "South Africa" or "South Sudan" or Uganda or Tanzania or Zambia or Zimbabwe or Benin or "Burkina Faso" or Burundi or Cameroon or "Central African Republic" or Chad or Congo or "Cote d'Ivoire" or "Ivory Coast" or "Democratic Republic of the Congo" or "Equatorial Guinea" or Gabon or Gambia or Ghana or Guinea or "Guinea-Bissau" or Liberia or Mali or Niger or Nigeria or Senegal or "Sierra Leone" or Togo or "sub-Saharan Africa") and (HIV or "Human Immunodeficiency Virus" or "HIV-1") and (incidence or prospective or cohort or longitudinal or panel or seroconver* or sero-conver*) and

("female sex workers" or "female sex worker" or "women who sell sex" or "woman who sells sex" or prostitut* or "sex-worker" or "sex work" or "sex-work" or FSW))

2. limit 1 to yr="1990 - 2022"

**EMBASE:**

1. ((Kigali or Soweto or Johannesburg or Durban or Port Elizabeth or Mpumalanga or Abidjan or Harare or Mombasa or Nairobi or Brazzaville or "Dar es Salaam" or "Cape Town" or "Addis Ababa" or Douala or Accra or Luanda or Lusaka or Conakry or Kampala or Maputo or Freetown or Bangui or Bosaso or Angola or Botswana or Eswatini or Swaziland or Ethiopia or Kenya or Lesotho or Malawi or Mozambique or Namibia or "South Africa" or "South Sudan" or Uganda or Tanzania or Zambia or Zimbabwe or Benin or "Burkina Faso" or Burundi or Cameroon or "Central African Republic" or Chad or Congo or "Cote d'Ivoire" or "Ivory Coast" or "Democratic Republic of the Congo" or "Equatorial Guinea" or Gabon or Gambia or Ghana or Guinea or "Guinea-Bissau" or Liberia or Mali or Niger or Nigeria or Senegal or "Sierra Leone" or Togo or "sub-Saharan Africa") and (HIV or "Human Immunodeficiency Virus" or "HIV-1") and (incidence or prospective or cohort or longitudinal or panel or seroconver* or sero-conver*) and ("female sex workers" or "female sex worker" or "women who sell sex" or "woman who sells sex" or prostitut* or "sex-worker" or "sex work" or "sex-work" or FSW))
2. limit 1 to yr="1990 - 2022"

**Google Scholar:** [first five pages of results screened]

"HIV incidence" in "female sex worker*" in sub-saharan Africa

**Supplementary text 2: Bayesian log-linear mixed-effects model description**

We modelled incidence rate ratios using a Bayesian mixed-effects regression model. The number of new HIV infections $Y_{it}$ observed in women engaging in sex work (WESW) in year $t$ in study $i$ followed a poisson distribution:

$$Y_{it} \sim Poisson(E[y_{it}])$$

Where $Y_{it}$ is observed new HIV infections in WESW and $E[y_{it}]$ is the expected number of infection events, which can be expressed as:

$$E[y_{it}] = \lambda_{it} * \mathrm{X}_{it}$$

Where$\lambda_{it}$ is the incidence rate in WESW and $X_{it}$ is the person years of follow-up. We expressed the incidence rate among WESW as the product of the matched total population female HIV incidence rate and an incidence rate ratio for WESW as follows:

$$\lambda_{it} = IRR_{it} * Z_{it}$$

such that, $\mathrm{IRR}_{it}$ is the incidence rate ratio and $\mathrm{Z}_{it}$ is the district-year-sex matched total population incidence. We model $\log(IRR)$ as:

$$\log(IRR) = \beta_0 + \beta_1 t + a_s + \mathrm{b}_s t$$

$$a_s \sim N(0, \sigma_a)$$

$$b_s \sim N(0, \sigma_b)$$

whereby $\beta_0$is the intercept, $\beta_1 t$ is a fixed effect for median-centred year, $\mathrm{a}_s$ represents study-level random intercepts, and $\mathrm{b}_s t$ captures study-level random slopes over time. Total population incidence and person-years of follow-up are used as model offsets.

$$\log\frac{1}{\sigma_a^2}, \log\frac{1}{\sigma_b^2} \sim N(1.6, 2)$$

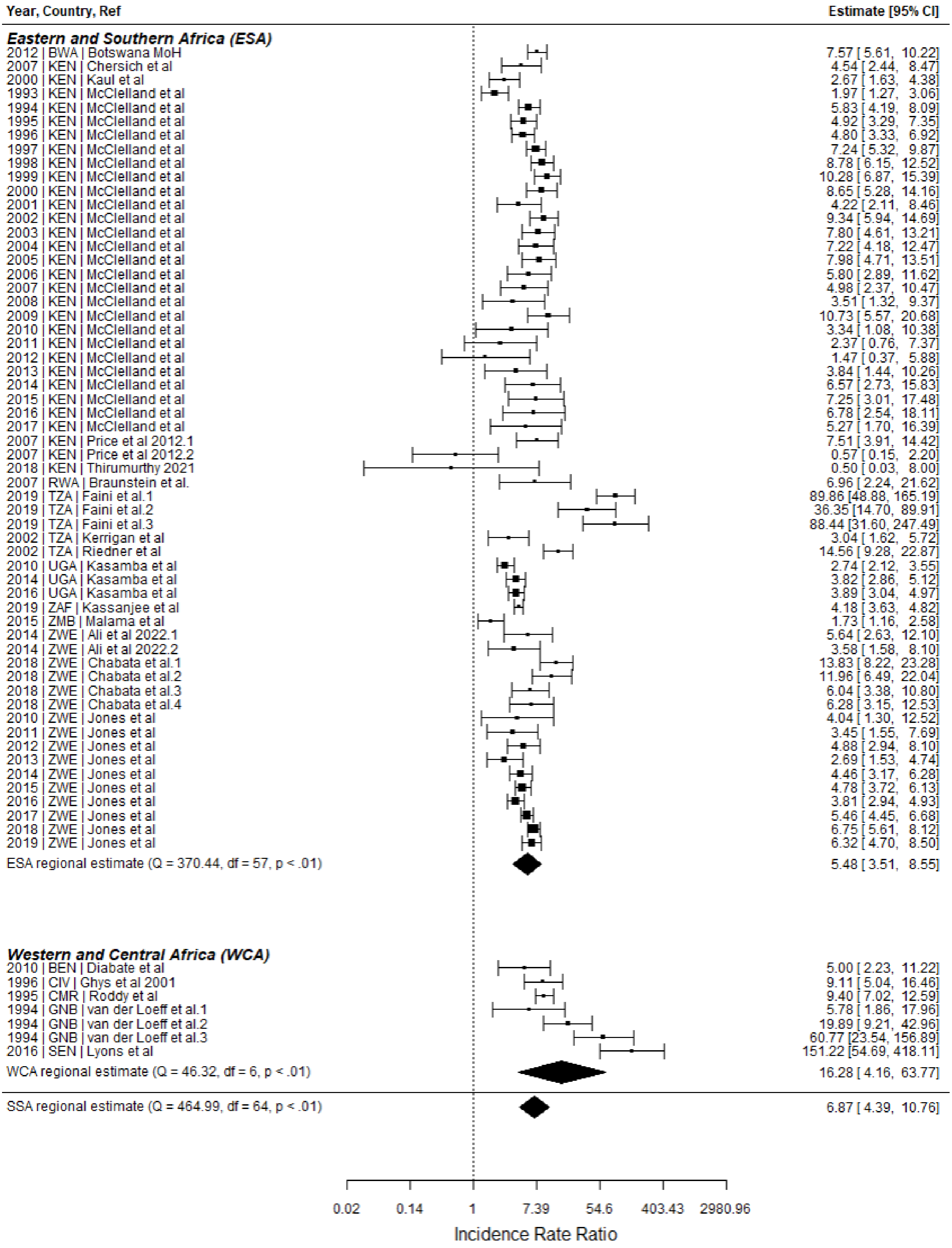

**Supplementary Figure 1: Meta-analysis of HIV incidence in women who engage in sex work relative to the total female population in Sub-Saharan Africa using only higher quality studies.** IRRs calculated by dividing empirical estimates of FSW HIV incidence by HIV incidence among district-age-year matched total population women derived from the Spectrum national estimates. Studies with a quality score lower than 60% excluded from this analysis. ESA: Eastern and Southern Africa; WCA: Western and Central Africa; SSA: Sub-Saharan Africa.

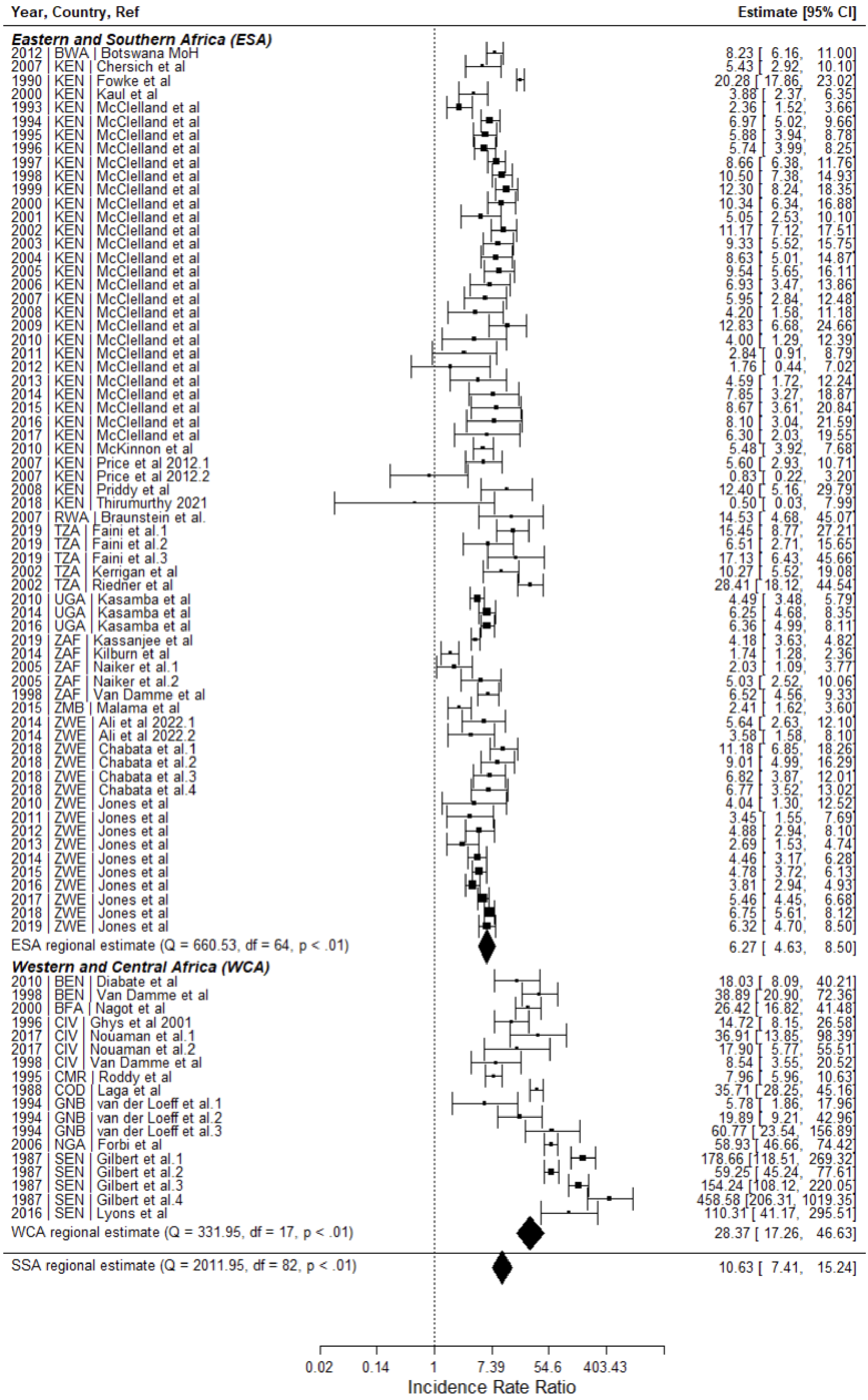

Year, Country, Ref
Estimate [95% CI]
Eastern and Southern Africa (ESA)
2012 | BWA | Botswana MoH 8.23 [ 6.16, 11.00]
2007 | KEN | Chersich et al 5.43 [ 2.92, 10.10]
1990 | KEN | Fowke et al 20.28 [ 17.86, 23.02]
2000 | KEN | Kaul et al 3.88 [ 2.37, 6.35]
1993 | KEN | McClelland et al 2.36 [ 1.52, 3.66]
1994 | KEN | McClelland et al 6.97 [ 5.02, 9.66]
1995 | KEN | McClelland et al 5.88 [ 3.94, 8.78]
1996 | KEN | McClelland et al 5.74 [ 3.99, 8.25]
1997 | KEN | McClelland et al 8.66 [ 6.38, 11.76]
1998 | KEN | McClelland et al 10.50 [ 7.38, 14.93]
1999 | KEN | McClelland et al 12.30 [ 8.24, 18.35]
2000 | KEN | McClelland et al 10.34 [ 6.34, 16.88]
2001 | KEN | McClelland et al 5.05 [ 2.53, 10.10]
2002 | KEN | McClelland et al 11.17 [ 7.12, 17.51]
2003 | KEN | McClelland et al 9.33 [ 5.52, 15.75]
2004 | KEN | McClelland et al 8.63 [ 5.01, 14.87]
2005 | KEN | McClelland et al 9.54 [ 5.65, 16.11]
2006 | KEN | McClelland et al 6.93 [ 3.47, 13.86]
2007 | KEN | McClelland et al 5.95 [ 2.84, 12.48]
2008 | KEN | McClelland et al 4.20 [ 1.58, 11.18]
2009 | KEN | McClelland et al 12.83 [ 6.68, 24.66]
2010 | KEN | McClelland et al 4.00 [ 1.29, 12.39]
2011 | KEN | McClelland et al 2.84 [ 0.91, 8.79]
2012 | KEN | McClelland et al 1.76 [ 0.44, 7.02]
2013 | KEN | McClelland et al 4.59 [ 1.72, 12.24]
2014 | KEN | McClelland et al 7.85 [ 3.27, 18.87]
2015 | KEN | McClelland et al 8.67 [ 3.61, 20.84]
2016 | KEN | McClelland et al 8.10 [ 3.04, 21.59]
2017 | KEN | McClelland et al 6.30 [ 2.03, 19.55]
2010 | KEN | McKinnon et al 5.48 [ 3.92, 7.68]
2007 | KEN | Price et al 2012.1 5.60 [ 2.93, 10.71]
2007 | KEN | Price et al 2012.2 0.83 [ 0.22, 3.20]
2008 | KEN | Priddy et al 12.40 [ 5.16, 29.79]
2018 | KEN | Thirumurthy 2021 0.50 [ 0.03, 7.99]
2007 | RWA | Braunstein et al. 14.53 [ 4.68, 45.07]
2019 | TZA | Faini et al.1 15.45 [ 8.77, 27.21]
2019 | TZA | Faini et al.2 6.51 [ 2.71, 15.65]
2019 | TZA | Faini et al.3 17.13 [ 6.43, 45.66]
2002 | TZA | Kerrigan et al 10.27 [ 5.52, 19.08]
2002 | TZA | Riedner et al 28.41 [ 18.12, 44.54]
2010 | UGA | Kasamba et al 4.49 [ 3.48, 5.79]
2014 | UGA | Kasamba et al 6.25 [ 4.68, 8.35]
2016 | UGA | Kasamba et al 6.36 [ 4.99, 8.11]
2019 | ZAF | Kassanjee et al 4.18 [ 3.63, 4.82]
2014 | ZAF | Kilburn et al 1.74 [ 1.28, 2.36]
2005 | ZAF | Naiker et al.1 2.03 [ 1.09, 3.77]
2005 | ZAF | Naiker et al.2 5.03 [ 2.52, 10.06]
1998 | ZAF | Van Damme et al 6.52 [ 4.56, 9.33]
2015 | ZMB | Malama et al 2.41 [ 1.62, 3.60]
2014 | ZWE | Ali et al 2022.1 5.64 [ 2.63, 12.10]
2014 | ZWE | Ali et al 2022.2 3.58 [ 1.58, 8.10]
2018 | ZWE | Chabata et al.1 11.18 [ 6.85, 18.26]
2018 | ZWE | Chabata et al.2 9.01 [ 4.99, 16.29]
2018 | ZWE | Chabata et al.3 6.82 [ 3.87, 12.01]
2018 | ZWE | Chabata et al.4 6.77 [ 3.52, 13.02]
2010 | ZWE | Jones et al 4.04 [ 1.30, 12.52]
2011 | ZWE | Jones et al 3.45 [ 1.55, 7.69]
2012 | ZWE | Jones et al 4.88 [ 2.94, 8.10]
2013 | ZWE | Jones et al 2.69 [ 1.53, 4.74]
2014 | ZWE | Jones et al 4.46 [ 3.17, 6.28]
2015 | ZWE | Jones et al 4.78 [ 3.72, 6.13]
2016 | ZWE | Jones et al 3.81 [ 2.94, 4.93]
2017 | ZWE | Jones et al 5.46 [ 4.45, 6.68]
2018 | ZWE | Jones et al 6.75 [ 5.61, 8.12]
2019 | ZWE | Jones et al 6.32 [ 4.70, 8.50]
ESA regional estimate (Q = 660.53, df = 64, p < .01) 6.27 [ 4.63, 8.50]
Western and Central Africa (WCA)
2010 | BEN | Diabate et al 18.03 [ 8.09, 40.21]
1998 | BEN | Van Damme et al 38.89 [ 20.90, 72.36]
2000 | BFA | Nagot et al 26.42 [ 16.82, 41.48]
1996 | CIV | Ghys et al 2001 14.72 [ 8.15, 26.58]
2017 | CIV | Nouaman et al.1 36.91 [ 13.85, 98.39]
2017 | CIV | Nouaman et al.2 17.90 [ 5.77, 55.51]
1998 | CIV | Van Damme et al 8.54 [ 3.55, 20.52]
1995 | CMR | Roddy et al 7.96 [ 5.96, 10.63]
1988 | COD | Laga et al 35.71 [ 28.25, 45.16]
1994 | GNB | van der Loeff et al.1 5.78 [ 1.86, 17.96]
1994 | GNB | van der Loeff et al.2 19.89 [ 9.21, 42.96]
1994 | GNB | van der Loeff et al.3 60.77 [ 23.54, 156.89]
2006 | NGA | Forbi et al 58.93 [ 46.66, 74.42]
1987 | SEN | Gilbert et al.1 178.66 [118.51, 269.32]
1987 | SEN | Gilbert et al.2 59.25 [ 45.24, 77.61]
1987 | SEN | Gilbert et al.3 154.24 [108.12, 220.05]
1987 | SEN | Gilbert et al.4 458.58 [206.31, 1019.35]
2016 | SEN | Lyons et al 110.31 [ 41.17, 295.51]
WCA regional estimate (Q = 331.95, df = 17, p < .01) 28.37 [ 17.26, 46.63]
SSA regional estimate (Q = 2011.95, df = 82, p < .01) 10.63 [ 7.41, 15.24]
0.02
0.14
1
7.39
54.6
403.43
Incidence Rate Ratio

**Supplementary Figure 2: Meta-analysis of HIV incidence in female sex workers (FSW) relative to the national-matched total female population in Sub-Saharan Africa.** IRRs calculated by dividing empirical estimates of FSW HIV incidence by HIV incidence among country-age-year matched total population women derived from the Spectrum national estimates. ESA: Eastern and Southern Africa; WCA: Western and Central Africa; SSA: Sub-Saharan Africa.

**Supplementary Table 1: Regression model summaries.**

| Covariate | All data, Mean (95% CI) | Kenya Data, Mean (95% CI) | Zimbabwe Data, Mean (95% CI) |
|---|---|---|---|
| **Intercept (Year = 2003)** | 1.743 (1.331, 2.160) | 1.971 (1.267, 2.666) | 0.516 (-0.329, 1.356) |
| **Year** | -0.001 (-0.069, 0.068) | 0.013 (-0.002, 0.028) | 0.095 (0.046, 0.145) |
| **Study Random Intercepts** | 2.460 (1.059, 4.925) | 1.195 (0.540, 2.170) | 8.266 (1.831, 23.715) |
| **Study Random Slopes with respect to year** | 46.177 (18.746, 93.644) | | |

**Supplementary Table 2: Global HIV Quality Assessment Tool Summary Table**

| STUDY | Research objective | Defined study population | Representative of the target population | Sample size | Proportion agreeing to participate reported | Participation >85%? | Representative sample of the source population | Incidence measurement | Statistical analysis | Sampling | Study length | Retention | Methods to address loss to follow-up | TOTAL (N) | TOTAL % |
|---|---|---|---|---|---|---|---|---|---|---|---|---|---|---|---|
| Ali (2020) ǂ | 1 | 1 | 1 | 0 | 0 | 0 | 1 | 1 | 1 | 1 | - | - | - | 7 | 70% |
| Botswana MoH (2013) ǂ | 1 | 1 | 1 | 1 | 0 | 0 | 1 | 1 | 1 | 1 | - | - | - | 8 | 80% |
| Braunstein (2011) | 1 | 1 | 1 | 1 | 0 | 0 | 1 | 1 | 1 | 0 | 1 | 1 | 0 | 9 | 69% |
| Chabata (2021) | 1 | 1 | 1 | 1 | 1 | 1 | 1 | 1 | 1 | 1 | 1 | 0 | 1 | 12 | 92% |
| Chersich (2014) | 1 | 1 | 0 | 0 | 1 | 1 | 1 | 1 | 1 | 1 | 1 | 1 | 0 | 10 | 77% |
| Diabete (2018) | 1 | 1 | 1 | 0 | 0 | 1 | 1 | 1 | 1 | 0 | 1 | 1 | 0 | 9 | 69% |
| Faini (2022) | 1 | 1 | 1 | 0 | 0 | 0 | 1 | 1 | 1 | 1 | 1 | 1 | 0 | 9 | 69% |
| Forbi (2011) ǂ | 1 | 1 | 0 | 0 | 0 | 0 | 0 | 1 | 1 | 0 | - | - | - | 4 | 40% |
| Fowke (1996) | 1 | 1 | 1 | 0 | 0 | 0 | 0 | 1 | 1 | 0 | 1 | 0 | 0 | 6 | 46% |
| Ghys (2001) | 1 | 1 | 1 | 0 | 1 | 0 | 1 | 1 | 1 | 0 | 1 | 0 | 0 | 8 | 62% |
| Gilbert (2003) | 1 | 1 | 1 | 0 | 0 | 0 | 1 | 1 | 1 | 0 | 1 | 0 | 0 | 7 | 54% |
| Jones (2023) | 1 | 1 | 1 | 0 | 1 | 0 | 1 | 1 | 1 | 0 | 1 | 0 | 0 | 8 | 62% |
| Kasamba (2019) | 1 | 1 | 1 | 0 | 0 | 0 | 1 | 1 | 1 | 1 | 1 | 1 | 1 | 10 | 77% |
| Kassanjee (2022) ǂ | 1 | 1 | 1 | 1 | 0 | 0 | 1 | 1 | 1 | 1 | - | - | - | 8 | 80% |
| Kaul (2004) | 1 | 1 | 1 | 1 | 1 | 0 | 0 | 1 | 1 | 0 | 1 | 1 | 0 | 9 | 69% |
| Kerrigan (2017) | 1 | 1 | 1 | 1 | 0 | 0 | 1 | 1 | 1 | 1 | 1 | 0 | 0 | 9 | 69% |
| Kilburn (2018) | 1 | 1 | 0 | 0 | 0 | 0 | 0 | 1 | 1 | 0 | 1 | 0 | 0 | 5 | 38% |
| Laga (1994) | 1 | 1 | 0 | 0 | 0 | 0 | 0 | 0 | 1 | 0 | 1 | 0 | 0 | 4 | 31% |
| Lyons (2020) | 1 | 1 | 1 | 0 | 0 | 0 | 1 | 1 | 1 | 1 | 1 | 0 | 0 | 8 | 62% |
| Malama (2022) | 1 | 1 | 1 | 0 | 1 | 0 | 1 | 1 | 1 | 1 | 1 | 0 | 0 | 9 | 69% |
| McClelland (2006) | 1 | 1 | 1 | 0 | 0 | 0 | 1 | 1 | 1 | 1 | 1 | 0 | 1 | 9 | 69% |
| McKinnon (2015) | 1 | 1 | 1 | 0 | 0 | 0 | 0 | 1 | 1 | 1 | 1 | 0 | 0 | 7 | 54% |

| | | | | | | | | | | | | | | | |
|---|---|---|---|---|---|---|---|---|---|---|---|---|---|---|---|
| Nagot (2005) | 1 | 1 | 0 | 0 | 0 | 0 | 0 | 1 | 1 | 0 | 1 | 1 | 0 | 6 | 46% |
| Naicker (2015) | 1 | 1 | 1 | 0 | 1 | 1 | 0 | 1 | 1 | 0 | 1 | 0 | 0 | 8 | 62% |
| Nouaman (2022) ǂ | 1 | 1 | 1 | 0 | 0 | 0 | 0 | 1 | 1 | 0 | - | - | - | 5 | 50% |
| Price (2012) | 1 | 1 | 1 | 0 | 1 | 1 | 1 | 1 | 1 | 1 | 1 | 1 | 0 | 11 | 85% |
| Priddy (2011) | 1 | 1 | 1 | 0 | 0 | 0 | 0 | 0 | 1 | 0 | 0 | 1 | 0 | 5 | 38% |
| Riedner (2006) | 1 | 1 | 1 | 0 | 0 | 0 | 0 | 1 | 1 | 0 | 1 | 1 | 1 | 8 | 62% |
| Roddy (1998) | 1 | 1 | 1 | 1 | 1 | 0 | 0 | 1 | 1 | 0 | 1 | 1 | 0 | 9 | 69% |
| Schim van der Loeff (2001) | 1 | 1 | 1 | 0 | 0 | 0 | 1 | 1 | 1 | 1 | 1 | 0 | 1 | 9 | 69% |
| Thirumurthy (2021) | 1 | 1 | 1 | 1 | 1 | 1 | 1 | 1 | 1 | 1 | 1 | 1 | 0 | 12 | 92% |
| Van Damme (2002) | 1 | 1 | 0 | 1 | 0 | 0 | 0 | 1 | 1 | 0 | 1 | 0 | 0 | 6 | 46% |
| TOTAL (N) | 32 | 32 | 26 | 9 | 10 | 6 | 19 | 30 | 32 | 15 | 26 | 12 | 5 | | |
| TOTAL (%) | 100% | 100% | 81% | 28% | 31% | 19% | 59% | 94% | 100% | 47% | 81% | 38% | 16% | | |
